# Generating synthetic data in digital pathology through diffusion models: a multifaceted approach to evaluation

**DOI:** 10.1101/2023.11.21.23298808

**Authors:** Matteo Pozzi, Shahryar Noei, Erich Robbi, Luca Cima, Monica Moroni, Enrico Munari, Evelin Torresani, Giuseppe Jurman

## Abstract

Synthetic data has recently risen as a new precious item in the computational pathologist’s toolbox, supporting several tasks such as helping with data scarcity or augmenting training set in deep learning. Nonetheless, the use of such novel resources requires a carefully planned construction and evaluation, to avoid pitfalls such as the generation of clinically meaningless artifacts.

As the major outcome described in the current manuscript, a novel full stack pipeline is introduced for the generation and evaluation of synthetic pathology data powered by a diffusion model. The workflow features, as characterizing elements, a new multifaceted evaluation strategy with an embedded explainability procedure effectively tackling two critical aspects of the use of synthetic data in health-related domains.

An ensemble-like strategy is adopted for the evaluation of the produced data, with the threefold aim of assessing the similarity of real and synthetic data through a set of well-established metrics, evaluating the practical usability of the generated images in deep learning models complemented by explainable AI methods, and validating their histopathological realism through a dedicated questionnaire answered by three professional pathologists.

The pipeline is demonstrated on the public GTEx dataset of 650 WSIs, including five different tissues, conditioning the training step of the underlying diffusion model. An equal number of tiles from each of these five tissues are then generated. Finally, the reliability of the generated data is assessed using the proposed evaluation pipeline, with encouraging results. We show that each of these evaluation steps are necessary as they provide complementary information on the generated data’s quality.

Overall, all the aforementioned features characterize the proposed workflow as a fully-fledged solution for generative AI in digital pathology representing a potentially useful tool for the digital pathology community in their transition towards digitalization and data-driven modeling.

## Introduction

In recent years, rapid advancements in scanning technologies have revolutionized traditional pathology, giving rise to the field of digital pathology (1) (2). The transition from conventional glass slides to high-resolution Whole-Slide Images (WSIs) has presented exciting opportunities for harnessing artificial intelligence, particularly deep learning, to revolutionize pathology, paving the way for computational pathology (1). Deep learning algorithms have demonstrated impressive capabilities in automating critical tasks such as cancer detection or grading (1) (3) (4) (5) (6) as well as cell segmentation (7) (8). However, the success of these deep learning algorithms heavily relies on the availability and quality of large-scale, annotated datasets (9), which can be challenging in the medical domain where data scarcity and privacy concerns are prominent (10). To address this issue, synthetic data generation has gained momentum, by producing data that maintains similar statistical properties as real data, making it a promising avenue to tackle privacy concerns while working with sensitive data such as medical images (11) (12).

At the heart of synthetic data generation lie generative models, such as Generative Adversarial Networks (GANs) (13). GANs have found widespread application in generating synthetic images across various medical domains (14) (15) (16), including digital histopathology images (17) (18) (19) (20). However, recent advancements have shown that diffusion models (21) have surpassed GANs in generating both natural and medical images (22) (23). Diffusion models are a class of generative models inspired by non-equilibrium thermodynamics. These models employ a two-step process to learn the data distribution to generate new data afterwards. In the first step, a Markov chain of diffusion steps is defined, where noise is progressively added to the input. Then, the model learns to reverse this process by generating novel data starting from the noisy inputs (24). The usage of diffusion models in medical image generation includes (but is not limited to) brain magnetic resonance images (25) (26), microscopic blood cells images (27), positron emission tomography heart images (28), and chest x-ray (29). A comprehensive review on the subject has been recently published (30).

The first application of diffusion models in generating histopathology images dates to (31), where the authors successfully employed denoising diffusion models to generate Hematoxylin and Eosin (H&E) tiles of three different subtypes of gliomas. Their results demonstrated that diffusion models outperform GANs for this task based on common quantitative evaluation metrics used in image generation quality assessments. Following their success, the authors in (32) also demonstrated the ability and superiority of diffusion models over GANs in generating colon histopathology images containing six different cell nucleus types and their corresponding masks. Additionally, authors in (33) used vision transformer-based diffusion models to generate colon histopathology images with higher quality than those generated using GANs.

Before exploiting generated data, it is crucial to evaluate them. However, the evaluation of generated data remains a topic of debate, with various studies adopting different approaches. The lack of a standardized evaluation pipeline to comprehensively assess generated data poses a major caveat. In this study, we aim to tackle this issue by introducing a general framework for generating and, more importantly, thoroughly evaluating synthetic pathology images from various viewpoints. We will illustrate how these multifaceted evaluations yield complementary insights that cannot be obtained through any single method alone.

The proposed synthetic pathology data generation and evaluation pipeline, as demonstrated in **Figure 1**, begins with the preprocessing stage, where tiles are extracted from WSIs.

**Figure 1.**
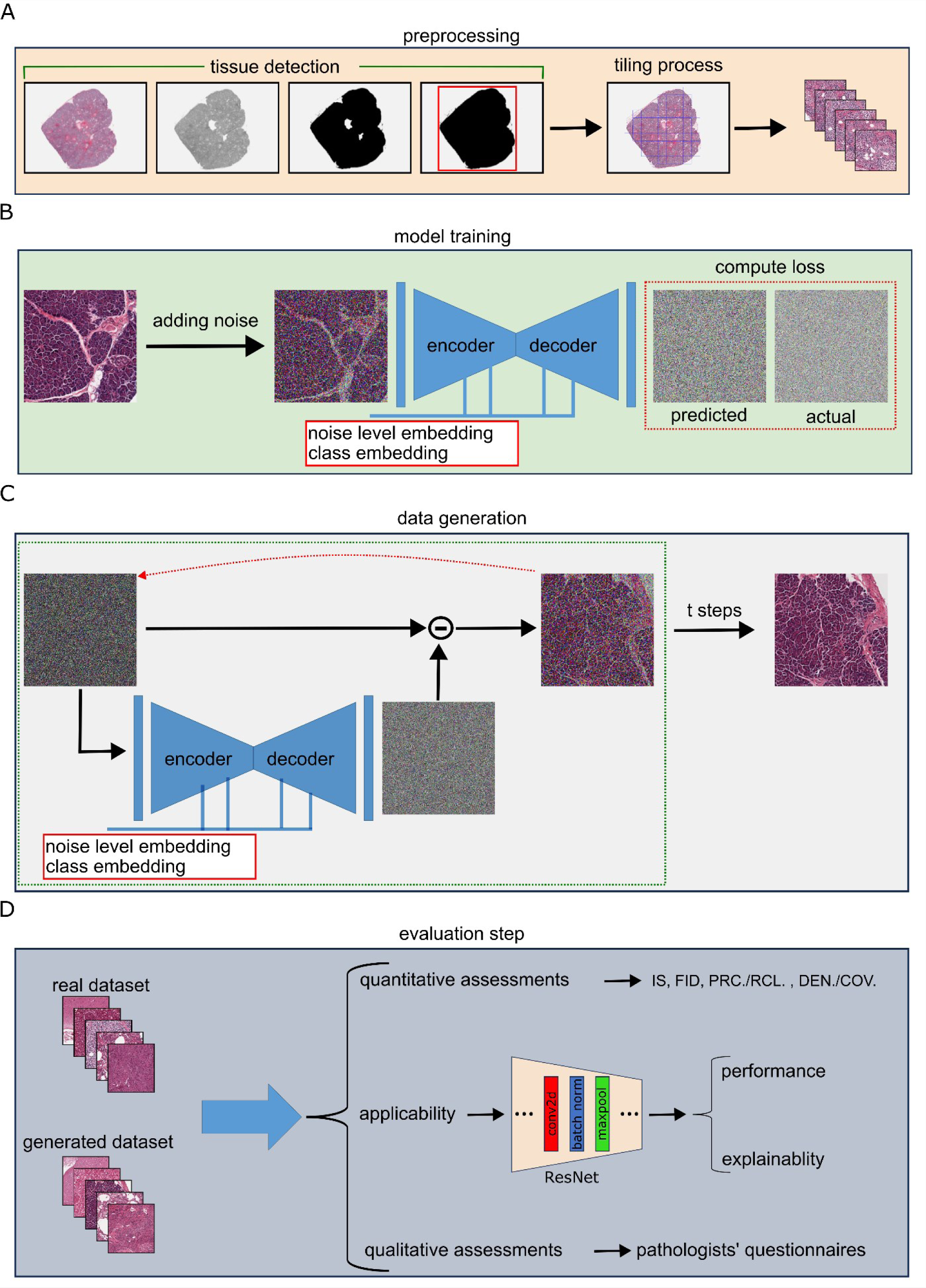
The proposed pipeline. The graphical workflow of our proposed pipeline is demonstrated. **(A)** Demonstrates the preprocessing step, where first, through a set of steps the tissue is detected in a slide followed by a tiling step to extract batches of tiles. (**B)** Demonstrates the model training step where the extracted tiles are input into a denoising diffusion model, where noise is initially added to images. These perturbed images, along with the noise level embedding and the class embedding (in our case, the tissue type), are subsequently fed into a U-Net model, which predicts the noise added to the image. Both the initial noise and the predicted noise are utilized to compute the loss and drive the training process. **(C)** Depicts the data generation step. Here, pure noise along with the desired class embedding are fed into the trained model to predict the noise image, which is then subtracted from the original noise to create a less noisy image. This resulting image serves as the new noisy input. After this process is repeated for ‘t’ steps, the final image is generated. **(D)** Subsequently, the generated dataset undergoes comprehensive evaluations, divided into three distinct categories. Quantitative assessment entails metrics such as Inception Score (IS), Frechet Inception Distance (FID), improved precision-recall (PRC, RCL), and density-coverage (DEN, COV) to gauge the similarity between the generated and real datasets. A practical evaluation involves training a ResNet classifier for tissue detection, assessing and comparing the usability of the generated data with the real dataset in terms of performance and explainability. Lastly, the biological realism of the generated data is evaluated through a series of questionnaires administered to expert pathologists.

Subsequently, a diffusion model is trained to generate data conditioned on different classes. After the training process, a sampling phase is performed to generate a specific number of synthetic data for each class. Lastly, the evaluation step process encompasses three distinct sets of assessments. The first set involves quantitative metrics that measure the similarity between the generated and real images. For our quantitative evaluation, we employed well-established metrics commonly used in the field of synthetic images, including Inception Score (IS) (34), Fréchet Inception Distance (FID) (35) improved precision-recall (36), and density-coverage (37).

The second set evaluates the practical usability of the generated images in deep learning models. To complement this set we investigated the convolutional filters learned by the classifiers, trained on both real and generated data, using the Concept Relevance Propagation (CRP) algorithm (38), an explainable artificial intelligence approach.

Lastly, a qualitative assessment is conducted to validate the biological realism of the generated data, a crucial aspect not targeted in previous assessments. This comprehensive approach ensures the reliability and usefulness of the generated dataset for our selected task. We applied the pipeline on 650 WSIs of five different tissues derived from the Genotype-Tissue Expression (GTEx) (39). We specifically chose to include multiple tissues, since most previous studies have focused on only one tissue type (31) (32) (33): such conservative approach fails to demonstrate whether similar success can be expected for other tissue types with varying complexities of morphology.

To the best of our knowledge, this study represents a novel and reproducible approach to histopathology image generation by covering multiple tissues and conducting a comprehensive evaluation of the generated data from various perspectives. By considering different tissues, and evaluation metrics, we aim to provide a more complete understanding of the capabilities and limitations of the proposed generative model. This multi-faceted evaluation allows us to gain insights into the effectiveness and applicability of the generated dataset for, potentially various, tasks.

## Materials and Methods

### Dataset

Images were obtained from the GTEx study (40). GTEx dataset contains WSIs collected using an Aperio glass slides scanner using H&E staining (39).

Among the 53 available human tissues, we retained data associated exclusively to samples obtained from our designated tissues of interest, namely the Brain, Kidney, Lung, Pancreas, and Uterus. This filtering process resulted in approximately 5000 samples’ metadata. Subsequently, we randomly selected a sample set of 650 specimens from the filtered metadata to obtain their corresponding WSIs. In cases where a WSI was not available for downloading, the specimen was excluded from the final dataset (the list of the downloaded WSIs is available in **Supplementary Table 1**).

Out of the 648 downloaded WSIs, a total of 589 have been used as training set for the deep learning models, while the remaining ones have been exclusively used for testing purposes.

### Preprocessing pipeline

WSIs demonstrate a considerably high spatial resolution: in our case, a spatial base resolution of 0.494 µm per pixel on a 20x magnification factor. Such granularity makes them impractical as direct inputs for neural networks.

It is commonly recommended to divide a WSI into uniform-sized tiles which can then be utilized as input for deep learning models (41) (42). Therefore, a preprocessing stage was deemed necessary to adequately prepare the collected data for model training.

In our study, we employed the Histolab^1^ library (43) to facilitate the tiling process of the slides. This library offers tools to generate tissue masks and perform the tiling process (43).

The WSIs do not necessarily contain only tissue information. Therefore, the creation of a tissue mask given a slide is a crucial step, as it ensures that the generated tiles primarily contain tissue-related information while minimizing extraneous background areas. We observed that the following ordered steps (depicted in **Figure 1A**) define an adequate tissue mask:

1. **RgbToGrayscale** filter: This process involves the conversion of a 3-channel image into a single channel grayscale image.
2. **OtsuThreshold** filter: Given a grayscale image, returns a binary thresholded image with separated foreground and background (44).
3. **BinaryDilation:** Performs a dilation on a binary image, i.e., the output image’s pixel value is determined by assigning it the maximum value among its neighboring pixels.
4. **BinaryFillHoles** filter: This process involves the task of filling all regions within a binary image.

In addition to the mask generation, other parameters are required in order to tile a WSI:

- **Tile Size:** Specifies the final resolution of the extracted tiles in pixels.
- **Level:** Specifies the level of tile extraction in a slide, the lower it is (minimum value 0), the higher the magnification factor will be.
- **Pixel Overlap:** Specifies in pixels how much the tiles will overlap between each other.

We decided to extract non-overlapping tiles with a resolution of 512×512 pixels from slides at the magnification factor of 5x (therefore with resolution of 1.976 µm per pixel). These parameters were chosen upon consultation with pathologists aiming to optimize the tissue classification task, providing sufficient contextual information for tissue differentiation, and avoiding excessive zooming that could render the tiles unidentifiable.

This resulted in 63247 tiles that were divided in two sets: from the 589 training WSIs, 56990 tiles have been extracted and used for training the generative model and the classifier, while the remaining 6257 tiles have been extracted from the 59 test WSIs and used for testing purposes. Nevertheless, even with a precisely defined mask, not all generated tiles may consistently capture informative attributes of the original slide. As illustrated in (the left panel of) **Supplementary Figure 1**, some tiles lack discernible morphological characteristics of the tissue, indicating that they may not contribute valuable information for the models. To filter such tiles, we exploited the *Complexity* metric (*C*) as inspired by (45) and it is defined as:

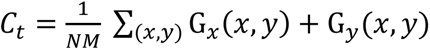

where the pixel-wise gradient (*G*) in the horizontal (*x*)and vertical (*y*) directions is computed for each tile *t*. The total sum of gradients is then multiplied by a term that is dependent on the width *N* and height *M* of the tile. A tile *t* with a non-uniform tissue will exhibit elevated pixel gradient values, thereby resulting in a correspondingly high value for *C*_*t*_.

We applied a chosen threshold (i.e., 0.4) based on visual inspection to retain only the tiles that exhibited sufficient distinguishable features. This step resulted in the removal of 948 uninformative tiles, allowing us to retain a total set of 62299 tiles, ranging from 10891 for Brain to 14173 for Pancreas. Additional details are summarized in **Supplementary Table 2**.

### Denoising Diffusion Probabilistic Model

For our generative model, we utilized the Denoising Diffusion Probabilistic Models (DDPM) proposed by (21). DDPM is a specific variant of diffusion models that focuses on denoising the input data by learning a probabilistic model capable of generating high-quality outputs. To create a dataset covering five different tissue types, we followed the classifier-free-guidance approach outlined in (22). In short, the pipeline works as follows.

First, Gaussian noise is introduced into the input images. The magnitude of this noise is determined randomly by a scheduler, resulting in varying levels of noise being applied to the input images. This approach allows the model to explore a spectrum of image noises, ranging from subtle distortions to an isotropic Gaussian distribution. The noisy image along with an embedding vector representing the level of noise and a trainable embedding vector representing the class (in our case the tissue type) are fed to a U-Net (46) based neural network, which aims to learn to output the noise. This prediction is then compared to the actual added noise, forming the basis for calculating the loss and guiding the model’s training process. This process is illustrated in **Figure 1B**.

Once the training phase is complete, the image generation phase begins. Starting with a baseline of pure isotropic noise, the trained network predicts the corresponding noise level. The difference between this predicted noise and the initial noise results in the creation of a less noisy image, which then becomes the input for the network once again. This iterative procedure is repeated for t steps, resulting in the generation of a final image, as illustrated in **Figure 1C**.

The model implementation can be found online^2^. The images were resized to 256 pixels, as experimenting with different sizes revealed that this produced the most favorable outcomes. For model training, we employed a batch size of 26. The learning rate was set at 1e-5, and the training procedure encompassed 215000 steps. We empirically found the learning rate by monitoring the sampled data after every 1000 steps. The number of steps for data generation (t) was set to 250. Visual inspection of image generation and model checkpointing was done every 1000 steps. Training was done till a convergence in the training loss was noticed as well as sampling during the training provided convincing results, since it has been observed that the quality of the generated images can be improved even when the training loss was stable. Class conditioning was done using a trainable torch.nn.Embeddings variable. Lastly, the generated images were further enhanced using a sharpening filter with a factor of 3.

### Evaluations

To comprehensively evaluate the quality of the generated images from various perspectives, we devised multiple sets of experiments ranging from widely recognized metrics for assessing generated images quality as well as expert human evaluation. In the following, we will give an overview of the details of each of these experiments.

### Canonical quantitative evaluation metrics

We assessed the quality of the generated images using well-established metrics in the field of synthetic images, including Inception Score (IS) (34), Fréchet Inception Distance (FID) (35), improved precision-recall (36), and density-coverage (37). Here, we will briefly explain each metric.

The IS is calculated using the following equation:

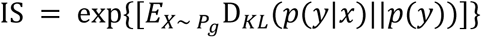

where *y* is a set of classes (in our case tissues), *x* is the image, *p*(*x*|*y*) is the conditional probability, *p*(*y*) is the marginal class probability, D_*KL*_ stands for the Kullback Leibler divergence and *C*_*x* ∼ *Pg*_denotes average over all the generated images.

High values of IS indicate that the generated images have a high diversity across different classes while maintain good within class similarities (34).We utilized the torchmetrics^3^ implementation to calculate IS in our study, splitting the images 5 times and using the Inception_v3 network (47) unbiased logits as the features to calculate the conditional and marginal probabilities.

Fréchet Inception Distance (FID) is a measure to quantify the similarity between the distributions of the generated and real images (35). It achieves so by passing the generated and real images through the original Inception_v3 network, extracting the outputs from a specified layer, and calculating the following formula:

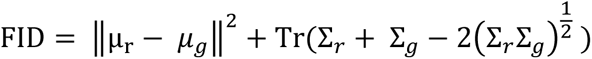

Here, *μ*_*r*_ and Σ_*r*_ represent the estimated average and covariance matrices of real images, while *μ*_*g*_ and Σ_*g*_ denote the same for generated images in the Inception\textunderscore{v3} feature space. The trace operation is denoted by Tr. To calculate FID, we utilized the torchmetric implementation, setting the feature parameter to 2048, which corresponds to the last pooling layer. Lower values of FID would indicate that the distributions of the real and generated images are more similar (35).

Both FID and IS rely on the imagenet (48) pre-trained Inception_v3 network which might not be ideal for those images whose distribution significantly differs from natural images (49) (50). To account for this, we additionally incorporated methods that assess the quality of generated images by examining the similarity of the manifolds between the real and generated data. The authors in (36) have proposed improved precision and recall measuring the overlap of the manifolds of the generated and real data. Briefly, they are calculated by projecting the data to a feature space and estimating the manifolds for real and generated data by surrounding each datapoint with a hyper-sphere that reaches its kth nearest neighbor (36). Precision is defined as the percentage of generated samples within the real data manifold, while recall is defined as the percentage of real images that fall within the generated data manifold (36). Higher values for both indicate greater overlap between the real and generated data. Precision and recall have been demonstrated to suffer in the presence of outliers (37). To overcome this problem, the authors in (37) have proposed the density and coverage metrics, which offer improved manifold estimation (for a more detailed explanation, please refer to (37)).

In this study, we calculated the precision, recall, density and coverage using the code provided by (37)^4^. We set the number of neighbors to 10. To prevent any potential bias that may rise due to networks being trained on specific datasets (37), the feature space for real and generated data was created by projecting the data onto a 100-dimension space through a ResNet-50 network (51) with random weights (for result comparability, we also performed analyses using an ImageNet pretrained network). We modified the last fully connected layer of the network to output a 100- dimensional vector. Since the imbalance between the number of real and generated images can bias the estimation (37), we addressed this issue by randomly sampling an equal number of generated images per tissue from the real data. To obtain reliable estimates, we repeated the entire process five times and reported the mean and standard error of the mean.

### Deep learning-based practical assessments

While the aforementioned evaluation measures provide valuable insights into the quality of generated images and their similarity to real ones, they may not provide sufficient information about more complex morphological features (such as histopathological plausibility) or the suitability of the generated images to be used as a replacement or in addition to the real images in machine learning studies. These aspects are of utmost importance and are one of the primaries focuses of our current study. Therefore, additional evaluation criteria are required. To do so, we designed a set of experiments using deep convolutional neural networks (CNNs) (52).

We used the ResNet-50 architecture (51) with randomly initialized weights to train a tissue classifier in two different scenarios. In the first scenario, we trained the network from scratch using the real tiles employed to train the diffusion model. Based on results demonstrated in (53), we put aside 30% of slides specifically for internal validation. This approach reduces the potential of data leakage related biases in the evaluation of models. To ensure that the network learns relevant morphological features associated with tissue types, we applied a set of following augmentations during training: spatial transformations, including random rotation and random horizontal-vertical flipping, chromatic transformations, such as jittering and random channel swapping, to mitigate the effects of staining (54), and robustness enhancing augmentations like blurring using a Gaussian filter and random erasing of parts of the image. The performance was evaluated on both the real tiles reserved for testing (check **Materials and Methods, Dataset** section) and the generated tiles.

The network was trained for 100 epochs with a batch size of 32. We employed the AdamW optimizer (55) with an initial learning rate of 0.0002. The learning rate was decreased using a cosine annealing scheduler with a tmax parameter of 100 (56). The model corresponding to the highest accuracy on the validation was saved for further steps. The model and training were implemented using pytorch^5^ and pytorchlightning^6^.

In the second scenario, adopting the exact same training setting as the first scenario, we trained a network from scratch using the generated tiles. The only distinction was that, since the generated images were on a tile level and not slide level, we reserved 30% of the tiles for internal validation. Subsequently, the trained network was evaluated on two distinct test sets. The first test set comprised a randomly selected 15% subset of the slides utilized in training of the diffusion model (referred to as the internal test-set). In contrast, the second test set consisted of real slides that were not seen by the diffusion model (the same test set used to assess the performance of the network trained on real data).

We evaluated the performance of the models on the test sets using multi-class accuracy (ACC) and Matthews’s correlation coefficient (MCC) (57) (58) (59).

### The explainability analysis

We employed Concept Relevant Propagation (CRP) (38) as a mean to assess whether the ResNet-50 utilized similar concepts, represented by convolution filters in CNNs, to classify images belonging to the same class, regardless of the dataset it was trained on. This analysis was crucial to ensure that the classifier employed consistent strategies in identifying the tissue of the tile, regardless of its origin (real or generated). The insights gained from this analysis provided valuable information for evaluating both the classifier itself and the quality of the synthetic tiles generated by the diffusion model.

CRP combines local and global approaches to provide robust and comprehensive explanations of a model’s predictions. It is based on Layer-wise Relevance Propagation (LRP) (60) but extended to disentangle explanations based on concepts learned by the network. CRP computes concept-conditional heatmaps to locate activation of concepts in the input space and provides dataset-level statistics to investigate how different classes or images activate specific channels in the model.

We extracted important concepts for each class from the network trained on the generated dataset by randomly selecting one generated image per class and choosing the convolutional layer 4 which, in the ResNet-50 architecture, is the last convolutional before the output. This choice is justified as we aimed to identify higher-level concepts, which are typically encoded in deeper layers closer to the output of the network (61) (62). The same process was done for the network trained on the real data.

Afterwards we constrained the relevance propagation by conditioning it on both the class and the layer. This allowed us to determine the significance of each channel in the selected layer for accurately classifying the given tile as belonging to its corresponding class.

In our analysis, we extracted a list of the top 5 channels per **class** per **model**, sorted by relevance. These channels were further utilized for feature visualization, where we obtained the most representative samples for each concept from both synthetic and real datasets, along with attribution heatmaps. Additionally, we assessed the prevalent class associated with these concepts, evaluating the integrity and efficacy of the concept extraction procedure. This analysis also helped in assessing the generalizability of the findings by examining whether the same associations hold true when employing the identical model on distinct datasets. This analysis was done using the CRP python package^7^.

### Qualitative evaluations

It is acknowledged that human expert evaluation remains the ultimate benchmark, particularly in the case of medical images. For this reason, we asked three professional pathologists (LC, EM, ET) to assist us in evaluating the quality of our generated images.

We did so by designing two different questionnaires. The first half of the first questionnaire included 100 questions. Pathologists were presented with three tiles from a single tissue and tasked with identifying the corresponding tissue. Half of the questions featured tiles derived from real data, while the remaining half consisted of generated images. The tiles “type” (real vs generated) was unknown to pathologists, the questions were randomized, and an equal number of tissues were included. The aim was to assess the pathologists’ ability to detect tissues using a limited number of tiles and to examine potential variations in performance between real and generated data. The second half of the first questionnaire consisted of an additional 100 questions. This time, pathologists were required to determine whether a given tile was real or generated, based on a single tile. The number of questions was equal for each tissue and data type (real or generated). Similar to the previous section, the questions were randomized across tissues and the distinction between real and generated data. The examples of the questions from the first questionnaire are depicted in **Supplementary Figure 2**.

The follow up questionnaire was sent one week after receiving responses to the first questionnaire. Pathologists were asked to rate from 0 (not important) to 5 (very important) how much they had employed some specific image features in distinguishing between real and generated tiles. The aim of this phase was to figure out whether the pathologists’ approach predominantly relied on “visual attributes”, such as blurriness, chromatic features, or the presence of artifacts, or if they also found biological or morphological irregularities in their discrimination between real and generated images. Out of the seven questions, four corresponded to visual attributes and three corresponded to biological attributes. The full list of questions is available in **Supplementary Table 3**.

### Implementation

All data processing and model training codes were implemented in Python (version 3.7.16 or higher). Any specific packages used were specified in the corresponding Materials and Methods section. For the evaluation questionnaires, we employed JavaScript and the Google Apps Script APIs.

The pre-processing, classifier training and evaluations were performed on a single machine with 2x Nvidia GTX-1080Ti GPUs, an Intel i7-6700 CPU, and 32GB RAM. The diffusion model training was conducted on a cluster with 2 x NVIDIA V-100 GPUs with 32GB VRAM.

All the codes are available in the Git-Hub repository {https://github.com/Mat-Po/diffusion_digital_pathology}.

## Results

We used the set of 56990 extracted real tiles as input for the diffusion model and performed training over 215000 steps. Confirming a converged loss and preliminary visual assessments of image quality, we produced 3001 synthetic images per tissue. This choice was motivated by our available resources and time constraints. A sample pair of real and generated images for each tissue are depicted in Figure 2.

**Figure 2.**
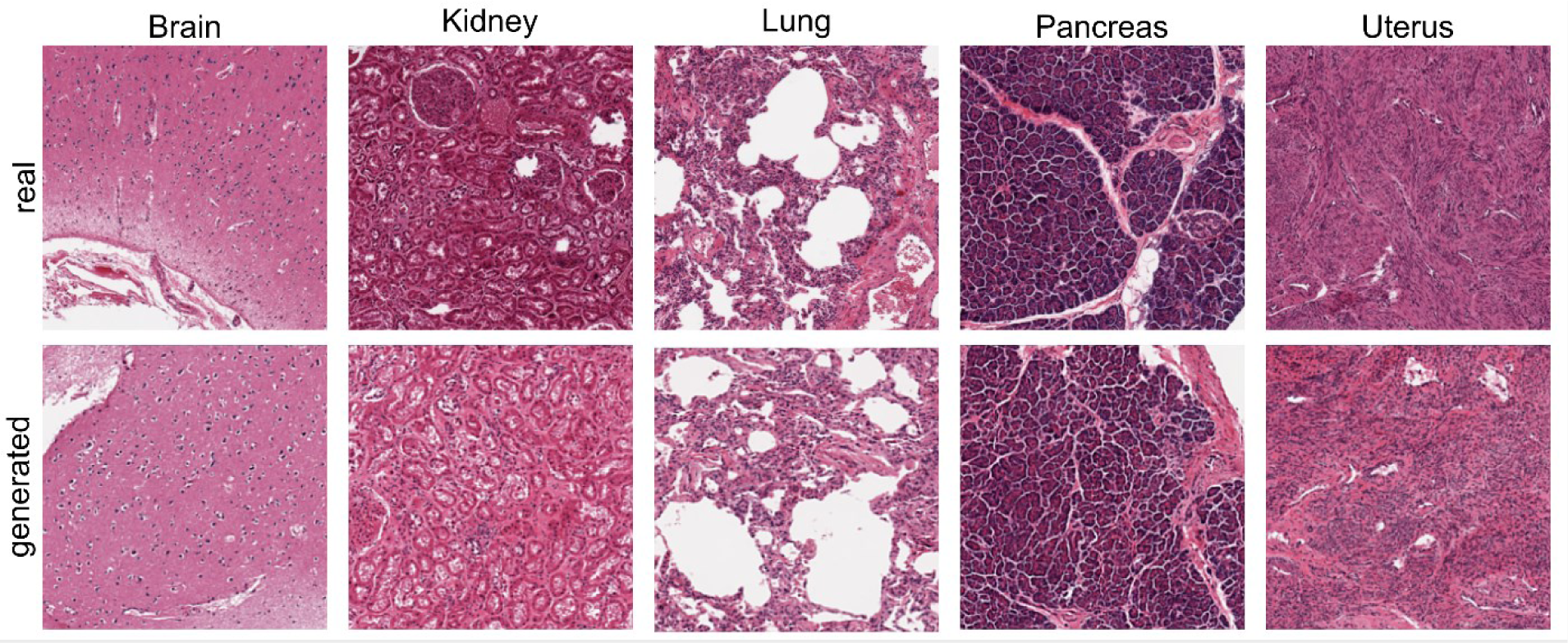
Example of real and generated image pair for each tissue. The top row displays a representative image from the real dataset for each tissue, while the bottom row showcases a generated image from the same tissue.

After our initial visual examinations, which confirmed a strong resemblance between the generated images and actual data, we moved forward with our evaluation pipeline.

### Measuring the Similarity Between Generated and Real Images Reveals Generated Data Resembles Real Images

After generating the synthetic dataset, we conducted a quantitative assessment of its quality using various synthetic image evaluation metrics. These results are summarized in **Table 1**.

**Table 1.**
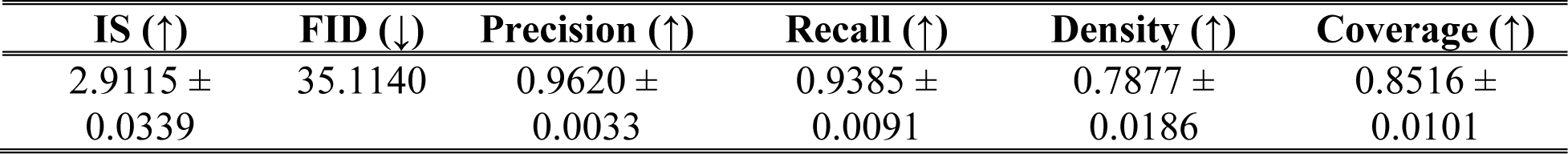
Results of quantitative evaluation metrics. The table presents the values for quantitative metrics, including the Inception Score (IS), precision, recall, density, coverage, and Fréchet Inception Distance (FID). The values for IS, precision, recall, density, and coverage are reported as the mean ± standard error of the mean based on repeated calculations. FID is calculated using the entire dataset. The arrows indicate whether higher (↑) or lower (↓) values are more desirable. Further details regarding the calculation and interpretation of these metrics can be found in the Materials and Methods section.

The overall results indicate that the generated images closely resemble the real images. Our generated images achieved a high Inception Score (2.9115 ± 0.0339), surpassing previous attempts at generating synthetic digital pathology images (31) (32) (33). In fact, the Inception Score achieved with our generated images is comparable to the score that would have been achieved with our real dataset (3.2892 ± 0.0149). The FID value, although slightly higher, is still comparable to values reported in other studies (31) (32) (33) (63). It is important to note that these studies utilized a smaller field of view, which can result in a lower FID. For instance, in (32), increasing the field of view by a factor of two (equivalent to our zoom level) more than doubled the FID (reaching a value of 38.1), which exceeds our reported FID. Additionally, based on results reported in (33), changing the dataset results in a significant change in the achieved FID. The precision and recall values approach the maximum value of 1 and outperform previous work (31) (33). However, it is important to consider that these studies calculated precision and recall using ImageNet pretrained networks, which can impact the results (37). To ensure a fairer comparison, we recalculated these values using an ImageNet pretrained ResNet50. The results demonstrate that our precision outperforms the other studies, while our recall remains comparable to their results. The summary of these comparisons can be found in **Supplementary Table 4**. To the best of our knowledge, no previous study has reported density and coverage metrics, but our values are also close to the maximum achievable value of 1, indicating a high similarity between the generated images and real ones. However, it is important to note that direct comparisons between studies may not be completely fair and feasible due to differences in datasets, tile sizes, zoom levels, and tissues. Our main focus in this paper is not solely on improving the quality of the generated images. Nevertheless, we present these quantitative metrics to demonstrate that our generated images exhibit similar quality to those of other previously published studies that have utilized diffusion models.

The calculated metrics provide an assessment of the overall quality of the generated dataset. However, for a more detailed analysis, we also calculated the FID individually for each class. The results are presented in **Table 2**. While an overall increase in FID was expected when calculating it separately for each class, we observed variations in this increase across different tissues. Specifically, the brain and pancreas tissues demonstrated a more substantial increase in FID compared to the uterus and lung tissues, which maintained their FID values close to the overall FID score. These results highlight the importance of examining the performance of the generative model at a class level, as it provides valuable insights into the variability of generated data quality across different tissue types.

**Table 2.** Class-wise FID. To have a better understanding of the quality of the generated data for each tissue, we calculated the Fréchet Inception Distance (FID) separately for each tissue type as described in the Materials and Methods. The FID scores are reported for each tissue class, indicating the dissimilarity between the generated and real images.

| **Brain** | **Kidney** | **Lung** | **Pancreas** | **Uterus** |
| --- | --- | --- | --- | --- |
| 58.92 | 45.89 | 39.19 | 64.98 | 33.97 |

### The Generated Data Exhibit Performance Comparable to Real Data in A Deep Learning-based Task

The performance of the trained ResNet-50 for tissue classification under various scenarios is summarized in **Table 3**. The table provides an overview of the test results obtained from different experimental settings. Furthermore, to gain a deeper understanding of the classification outcomes, the corresponding confusion matrices are visualized in **Supplementary Figure 3**.

**Table 3.** Performance of the trained neural networks for the tissue classification task. The accuracy (ACC) and Matthews correlation coefficient (MCC) values are reported for various training and testing scenarios as described in the Materials and Methods section.

| **Train set – Test set** | **ACC** | **MCC** |
| --- | --- | --- |
| Real data – External real data | 98% | 0.9705 |
| Real data – Generated data | 98% | 0.9796 |
| Generated data – Internal real data | 95% | 0.9331 |
| Generated data – External real data | 93% | 0.9133 |

The results indicate that there is no significant difference in the performance of the network trained on real data when tested on either unseen real data or generated data. This finding suggests that the generated images have successfully captured the diverse morphological patterns present in the original tissues, as the network’s ability to classify them remains consistent. In contrast, when evaluating the network trained on generated data, a decrease in performance is observed for both seen and unseen real data. It is noteworthy that earlier studies have also documented a reduction in performance when relying solely on generated data as opposed to real data (33). This reduction in performance can be attributed to several factors. Firstly, the smaller number of training samples available for the network trained on generated data compared to real data may contribute to lower accuracy. In fact, when we conducted an experiment by retraining the network using an equal number of real image tiles as generated ones, we observed a decrease in accuracy of approximately 1.5% (data not shown). Additionally, as evident from the confusion matrices, the decline in performance is notably pronounced for certain tissues such as pancreas and lung. This observation suggests that these tissues present more intricate and complex characteristics, potentially posing challenges for the diffusion model in generating diverse and accurate tiles. Interestingly, this phenomenon contrasts with our quantitative metrics, such as the class-wise FID, which might have predicted greater difficulties in tissues like the brain. However, the outcome of the deep learning assessment did not align with this expectation, underscoring the significance of multifaceted evaluations.

Despite these drawbacks, the overall results are promising, as they demonstrate that the generated images exhibit similar patterns to those found in real tiles. Furthermore, by utilizing a small number of generated images, we can achieve performance levels comparable to those attained using real data.

### Explainability-mediated analysis further reveals insights about the features learned from both datasets

We employed CRP to identify the top 5 channels or concepts activated for each class in both ResNets, one trained on real data and the other on generated data. These lists are presented in **Supplementary Table 5**. Once we determined the class-specific concepts, we used them to visualize which input pixels activate these concepts.

For instance, in **Figure 3A**, channel 428, associated with kidney-related concept in the ResNet trained on real data, is activated by kidney samples from both generated and real data. This is somewhat expected from the real data, given how we extracted this channel. However, it is not taken for granted for the generated data. Notably, the input pixels activating channel 428 are part of the same anatomical structure, the kidney’s glomerulus. In simpler terms, channel 428 in the ResNet trained on real data is activated by the glomeruli in kidney samples both in real and generated data.

**Figure 3.**
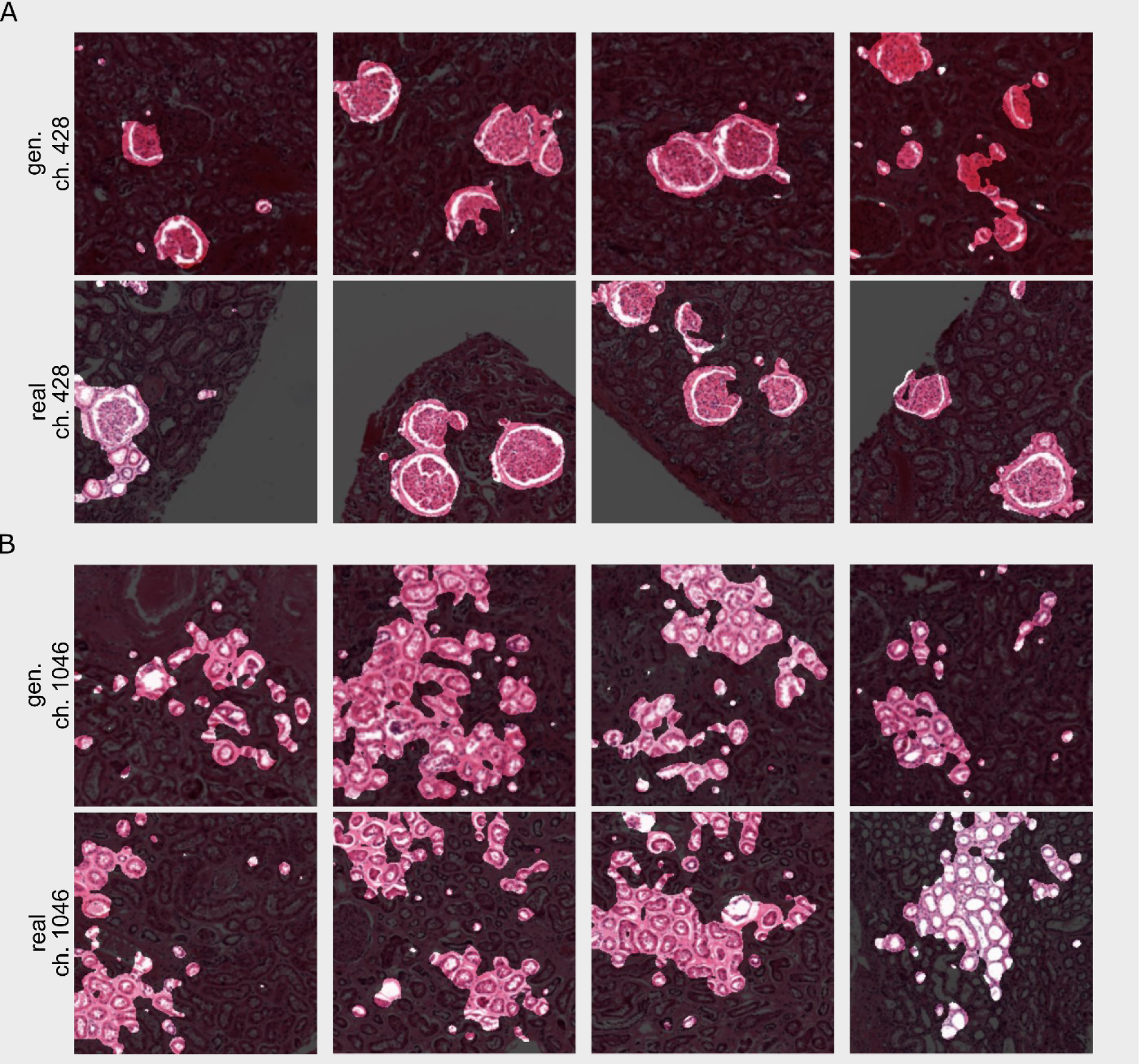
Concept relevance propagation visualization. Visualization of input pixels that activate the indicated channels. Each panel explores one network, either trained on real or synthetic data, and one concept. Each row examines the relevance of the channel either on the synthetic dataset (gen.) or on the real one (real). **(A)** ResNet trained on the real dataset. Input pixels that activate channel 428 in the last convolutional layer belong to the same tissue class and consistent morphological elements in both datasets. (**B)** ResNet trained on the synthetic dataset. Here, too, the tiles that activate the channel belong to the same class in both datasets, and input pixels belong to consistent morphological structures.

What’s crucial is that when we examine the concept that is most activated by kidney samples in the network trained on generated data, shown in **Figure 3B**, a similar pattern emerges. In both real and generated data, the input pixels activating this channel correspond to the glomeruli of kidney images.

We also used the global approach in CRP to identify which classes are most activated by the same set of selected channels. The upper part of **Supplementary Table 5** contains channels from the network trained on real images, while the lower part contains channels from the network trained on generated tiles. Upon inspecting the classes activated by each channel in both datasets, we found a strong agreement, suggesting that the networks behave similarly when it comes to tiles from both datasets. This evidence supports the performance metrics used to analyze the classifier’s performance.

Additionally, these results shed light on situations where the network makes mistakes, particularly in cases of confusion between pancreas and lung. Channel 890 from the lower part of **Supplementary Table 5** illustrates this, as although the most frequent class activated by that channel is Pancreas, the second most frequent class, with a substantial normalized relevance of 0.95 and 0.94 depending on the dataset, is the Lung class.

To gain a better understanding, we selected the most relevant concept for each tissue and visualized the pixels activating that concept in some example real and generated images using both the networks trained on the real dataset (**Supplementary Figure 4**) and the network trained on the generated dataset (**Supplementary Figure 5**). While it appears that the most important concept for tissues such as brain, kidney, lung, and uterus, to some extent, remains consistent between the networks trained on real and generated data, this isn’t the case for pancreas. For the network trained on generated data, the most important concept for pancreas tissue, while descriptive enough (as indicated by the classifier’s overall good performance), seems to resemble the most important concept for lung and uterus tissue, which explains the decrease in performance.

### The Generated Data is Biologically Realistic in the Eyes of Experts

The pathologists’ questionnaire results are summarized in **Table 4** and depicted in **Figure 4**.

**Figure 4.**
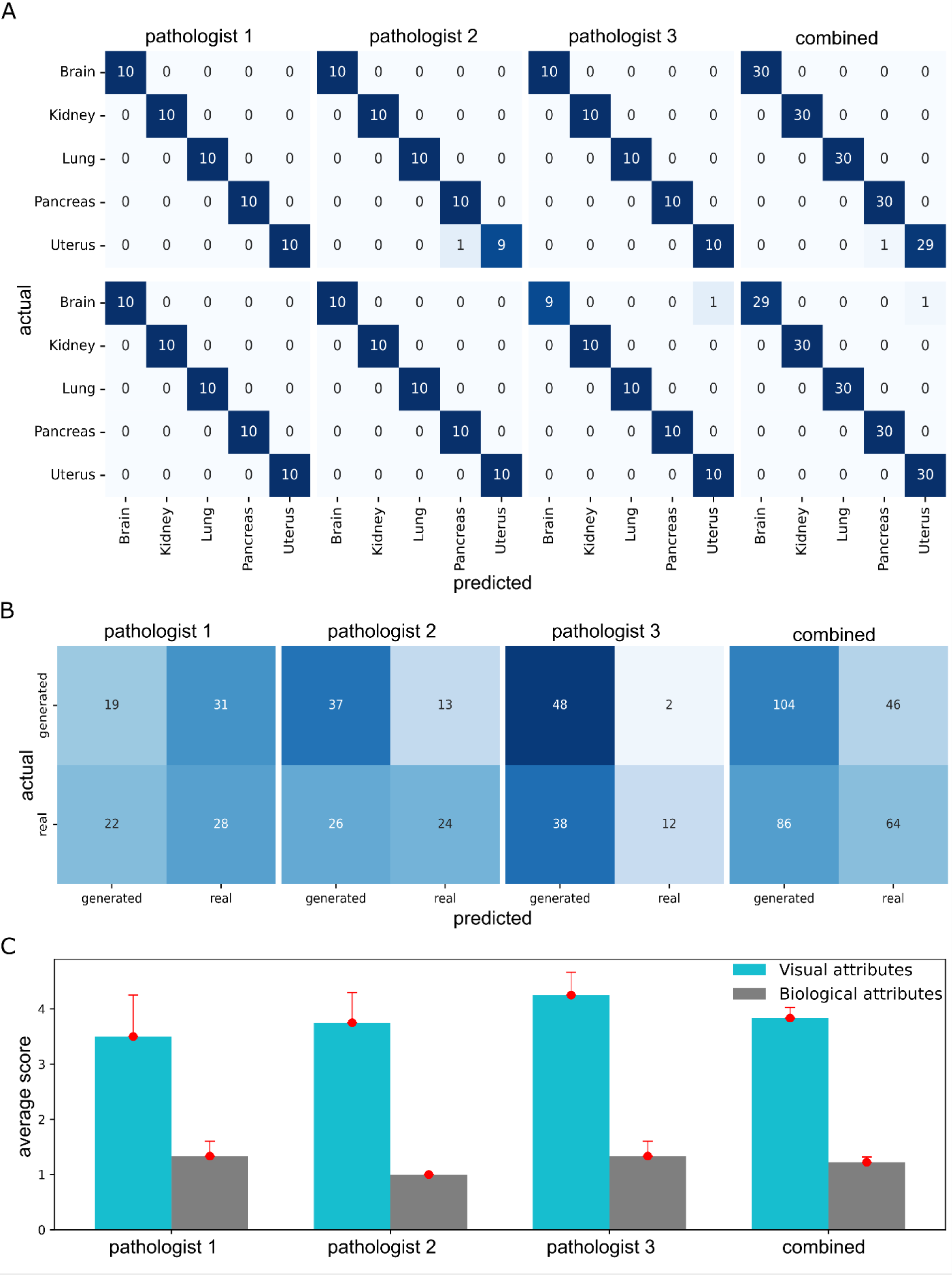
Pathologists’ questionnaire results. **(A)**Presents the confusion matrices illustrating the pathologists’ performance in tissue detection task in the first questionnaire, using both real (top row) and generated (bottom row) images. These matrices correspond to the first two rows of **Table 3**. (**B)** Displays the confusion matrices that depict the pathologists’ performance in the task of discriminating between real and generated images during the first questionnaire. These matrices correspond to the last row of the results presented in **Table 3**. **(C)** Illustrates the average ± standard error of the mean of the pathologists’ responses to the second questionnaire. The cyan bar represents the average score indicating the score to which pathologists relied on visual attributes to differentiate real images from generated ones, while the gray bar indicates the extent to which they utilized biological attributes as described in Materials and Methods.

**Table 4.** First questionnaire results. The accuracy (ACC) and Matthews correlation coefficient (MCC) values are reported for the tissue detection task using real and generated data, as well as for the real vs. generated discrimination task. The results are presented individually for each pathologist, and the combined results are obtained by aggregating the responses of all pathologists.

| *Task* | Pathologist 1 |  | Pathologist 2 |  | Pathologist 3 |  | Combined |  |
| --- | --- | --- | --- | --- | --- | --- | --- | --- |
|  | ACC | MCC | ACC | MCC | ACC | MCC | ACC | MCC |
| Tissue detection with real images | 100% | 1 | 98% | 0.9757 | 100% | 1 | 98.33% | 0.9917 |
| Tissue detection with generated images | 100% | 1 | 100% | 1 | 98% | 0.9757 | 98.33% | 0.9917 |
| Real or generated | 47% | -0.061 | 61% | 0.2278 | 60% | 0.2881 | 56% | 0.1245 |

The results from the first task of tissue detection demonstrate that pathologists are highly proficient in identifying the tissue type based on the given three tiles at the provided zoom level. Importantly, there is no significant difference in performance between real and generated images, indicating that the generated data accurately capture the morphology of the original tissues. The combined performance of the pathologists slightly exceeds chance level in detecting real vs. generated tiles (accuracy of 56%), suggesting that the generated tiles possess a biologically realistic appearance. When we examined the distribution of errors made for the generated tiles (generated tiles mistaken for real), as shown in **Supplementary Figure 6**, the brain images had slightly higher average number of mistakes, suggesting that they appeared more realistic to the pathologists’ eyes and conversely, kidney images had the lowest average number of mistakes, indicating that pathologists found it easier to detect generated kidney images. Nevertheless, the overall differences across tissues were small and not significant. Notably, the results from the second questionnaire reveal that the pathologists primarily relied on visual attributes such as chromatic features and blurriness to determine whether a tile was real or generated, as they did not detect any noticeable biological irregularities in the generated images. When combined, the average score given to visual attributes was 3.83 ± 0.19, which was found to be statistically significantly different from the average score of 1.22 ± 0.01 given to biological attributes (two-tailed paired t-test, p-value = 0.0075). This finding highlights the potential value of the generated dataset for educational purposes, as it indicates that the generated images are biologically realistic to the extent that even expert pathologists are almost unable to distinguish them, except for some minor differences at the image visual quality level.

## Discussion

The field of digital pathology, like many other disciplines, could greatly benefit from the ability to generate realistic synthetic data. These data could assist in addressing privacy concerns and enabling AI-assisted diagnosis by providing high-quality data to aid the training process. This holds promise in scenarios requiring pixel/tile-level annotations which can be time-consuming and susceptible to errors when executed by humans. Hence, in this article, we have introduced a comprehensive pipeline for generating and assessing synthetic data in digital pathology. We employed this pipeline to generate images from five distinct tissues with a wide field of view, employing a denoising diffusion model. The quality of the data was assessed through three distinct evaluation approaches: quantitative assessment employing well-established metrics, practical applicability evaluation through deep learning-based tissue classification and explainability, and qualitative evaluation involving questionnaires administered to pathologists. Our findings emphasize the fact that employing these varied evaluations provides us with complementary insights into data quality, highlighting that relying solely on a single approach would be insufficient.

Given the current trend towards analyzing digital pathology images at the whole slide level (41) (64) (65), it is desirable to generate synthetic data at the same level as well. In our study, we focused on generating tiles of wide field of view, to examine the ability of diffusion models in generating images at this resolution and as an early attempt to get closer to WSI image generation. Future research can build upon this by generating tiles from multiple fields of view and subsequently integrating them to generate complete whole slide images. Additionally, incorporating the adjacency of tiles in the generative model could assist in generating neighboring tiles to be able to generate WSIs. An example of this has been done with GANs in (66).

We explored the diffusion model’s capability to generate tiles from various tissues, each characterized by distinct morphological complexities. Our findings indicated that, although the overall performance was satisfactory, the model’s effectiveness varied across different tissues. An intriguing observation highlighting the importance of diverse evaluations emerged when examining each tissue individually.

In particular, when comparing class-wise FID scores, brain and pancreas showed similar results, both significantly higher than lung. However, classifier performance was notably lower for pancreas and lung, while brain exhibited almost perfect performance. Furthermore, the results from the pathologists’ questionnaire indicated that pathologists had the most difficulty in distinguishing between real and generated brain images, implying that the generated brain images are of the highest quality.

Explainability analyses shed further light on these findings. They revealed that the most relevant histopathological features for detecting pancreas when the network is trained on generated data differ from that when trained on real data. Interestingly, these features closely resemble the most relevant concepts for lung tissue. It remains to be determined whether these differences are attributed to the limitations of the diffusion model’s classifier-free guidance pipeline in distinguishing between lung and pancreas tissues or whether they stem from inherent complexities within the pancreas tissue itself.

Despite tissue classification not ranking among the foremost priorities in digital pathology, we harnessed its utility to assess the quality of generated data and their viability in replacing real tiles within deep learning-based tasks. As anticipated, the performance of a model trained solely on generated data did not match that of a network trained on real data, yet the outcome remained promising and satisfactory. The generation of data to facilitate network training becomes particularly valuable when dealing with scenarios that require pixel-level annotations, which are labor-intensive. Recent studies have showcased generative models’ potential in producing annotated pairs of tissue images alongside their corresponding cell annotations’ mask (32) (67) for specific tissues. This lays the foundation for expanding our work, potentially involving the generation of data from various tissues, accompanied by their respective cell type annotations under normal and various pathological conditions unique to each tissue.

Explainable artificial intelligence is a rapidly growing sub field within the machine learning community (68). It has emerged as a response to the challenge of interpreting predictions made by black box models, where the underlying mathematical functions are often highly complex and difficult to understand, even for AI experts. Deep neural networks, although highly effective in their performance when dealing with unstructured data such as images, fall into this category of black box models. To leverage the power of deep learning methods while addressing the need for interpretability, post-hoc methods have been developed. In this study, we demonstrated the application of an explainability technique to enhance our understanding of data quality and to gauge whether, for the generated data, the model learns comparable features as when trained with real data. This holds a special significance, especially considering a recent study that highlighted how generated histopathological images can provide explanations beyond what is attainable with real data (69). This is particularly important in the medical domain, where the demand for interpretable models remains crucial.

Finally, the pathologists’ relatively low performance in distinguishing between real and generated images reinforced our confidence in the realism of the generated images, thus promoting their utility for educational purposes. Notably, the feedback garnered from the subsequent questionnaire was intriguing. It emphasized that instances where pathologists successfully identified generated tiles were often attributable to discernible visual cues like blurriness or chromatic characteristics. Given the significantly high resolution of histopathology images, the fact that generated images would not consistently match this level of resolution was to be expected. This limitation could potentially be addressed in future studies by integrating super-resolution networks such as the one proposed in (70), to enhance the quality of generated images.

Overall, although synthetic data in digital pathology may not be ready to completely complement real images for research purposes, our findings, like other studies, highlight their promising potential in the future. What’s important to consider is the need to examine these generated data from different angles to fully understand their strengths and weaknesses. Our results emphasize the importance of thoroughly evaluating the generated dataset using various approaches, which should become a common practice in the field to better develop and use synthetic datasets effectively.

## Data Availability

All synthetic data produced in the present study are available upon reasonable request to the authors.
Code and model's weights are available online

https://github.com/Mat-Po/diffusion_digital_pathology

https://huggingface.co/pozzi/diffusion_digital_path/tree/main

## Fundings

The study was partially funded under the National Plan for Complementary Investments to the NRRP, project “D34H—Digital Driven Diagnostics, prognostics and therapeutics for sustainable Health care” (project code: PNC0000001), Spoke 2: “Multilayer platform to support the generation of the Patients’ Digital Twin”, CUP: B53C22006170001, funded by the Italian Ministry of University and Research.

## Data and Code Availability

All the data used in this paper is publicly accessible from https://gtexportal.org/home/. The source code is publicly available on GitHub at: https://github.com/Mat-Po/diffusion_digital_pathology ResNet and Diffusion model weights are publicly available on HuggingFace model hub at: https://huggingface.co/pozzi/diffusion_digital_path/tree/main

## Author Contribution

The study was conceived by GJ, who, along with SN, supervised the project. GJ secured funding. MP conceived the initial design. MP, SN, MM, LC, and GJ contributed to the final design and proposal of the methodology. ER conducted preprocessing of the slides and generated the questionnaires. MP performed diffusion model training and generated synthetic data, as well as explainability analyses. SN carried out quantitative evaluations and deep learning-based assessments. LC, EM, and ET participated in the quality assessment of the data and answered the questionnaires. The manuscript was written by SN, MP, ER, and GJ, with input from MM, LC, EM, and ET.

## Declaration of generative AI in scientific writing

During the preparation of this work the authors used ChatGPT3.5^8^ for grammar and spelling checks, as well as rephrasing to enhance readability. After using this tool/service, the authors reviewed and edited the content as needed and take full responsibility for the content of the publication.

## Competing Interest Statement

The authors declare no competing interests.

## Supplementary Information for

**Supplementary Figure 1.**
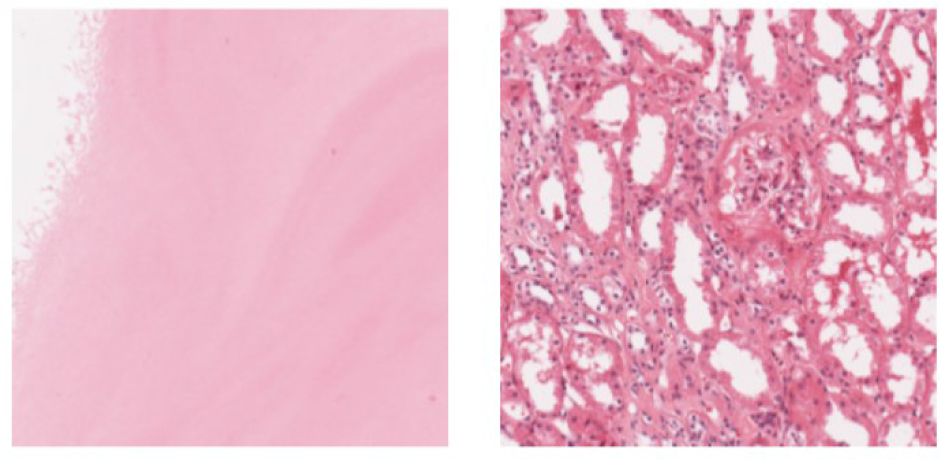
Differentiating non-informative and informative tiles. The left panel showcases a tile that lacks discernible tissue information, unlike the tile displayed in the right panel, which exhibits clear tissue characteristics. The complexity score for the left tile is 0.13, falling below our predetermined threshold, while the right tile scores 1.81 in complexity. Consequently, the non-informative left tile is excluded from further analyses, while the informative right tile is retained for subsequent investigations.

**Supplementary Figure 2.**
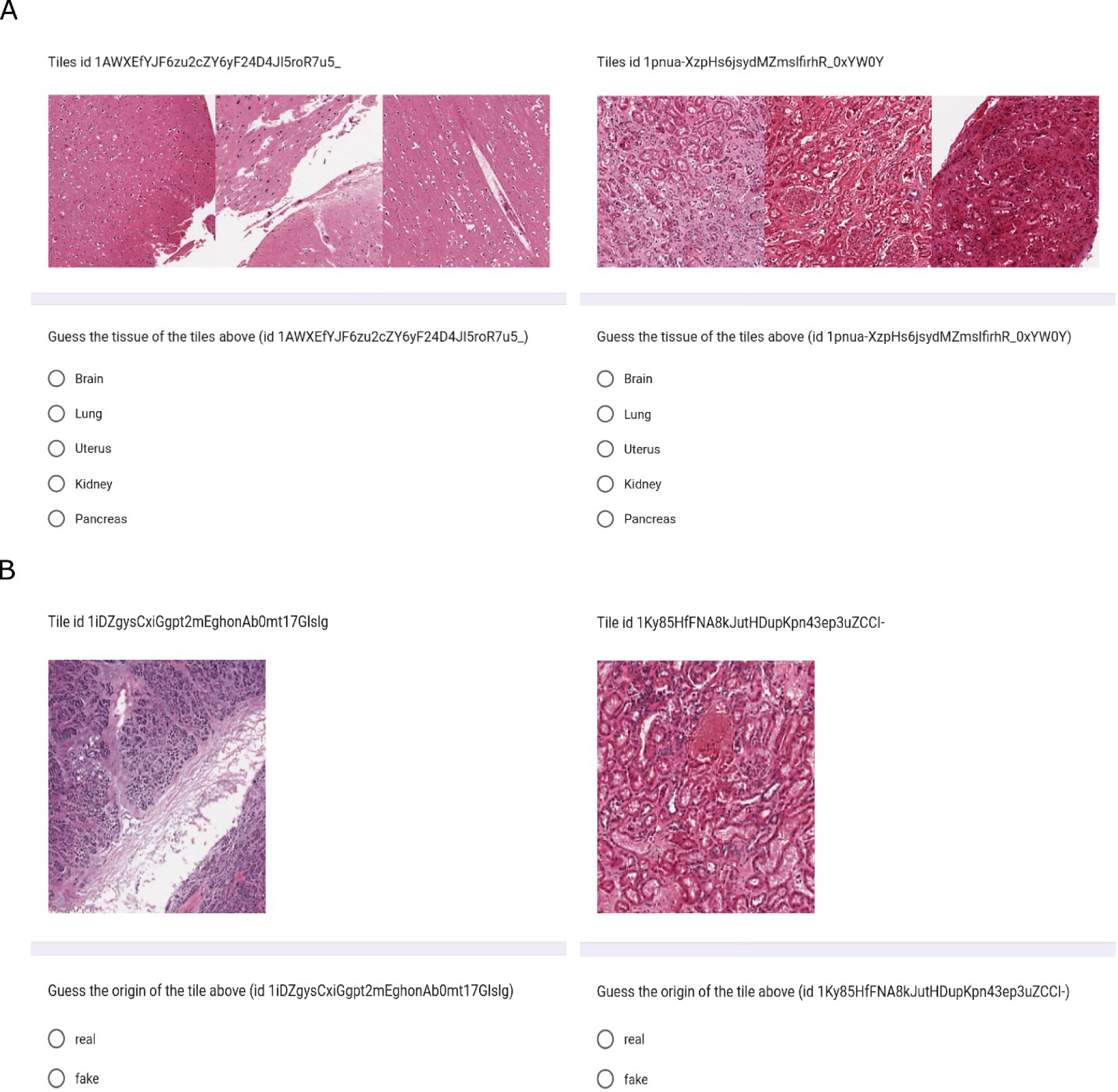
Example questions from the first questionnaire. **(A)** displays two example questions from the tissue detection task of the first questionnaire. The left image represents tiles extracted from real brain images, while the right image presents tiles from a generated kidney image. **(B)** illustrates two example questions from the real vs generated task of the first questionnaire. In the left image, a real pancreas image is shown, while the right image exhibits a generated kidney tile.

**Supplementary Figure 3.**
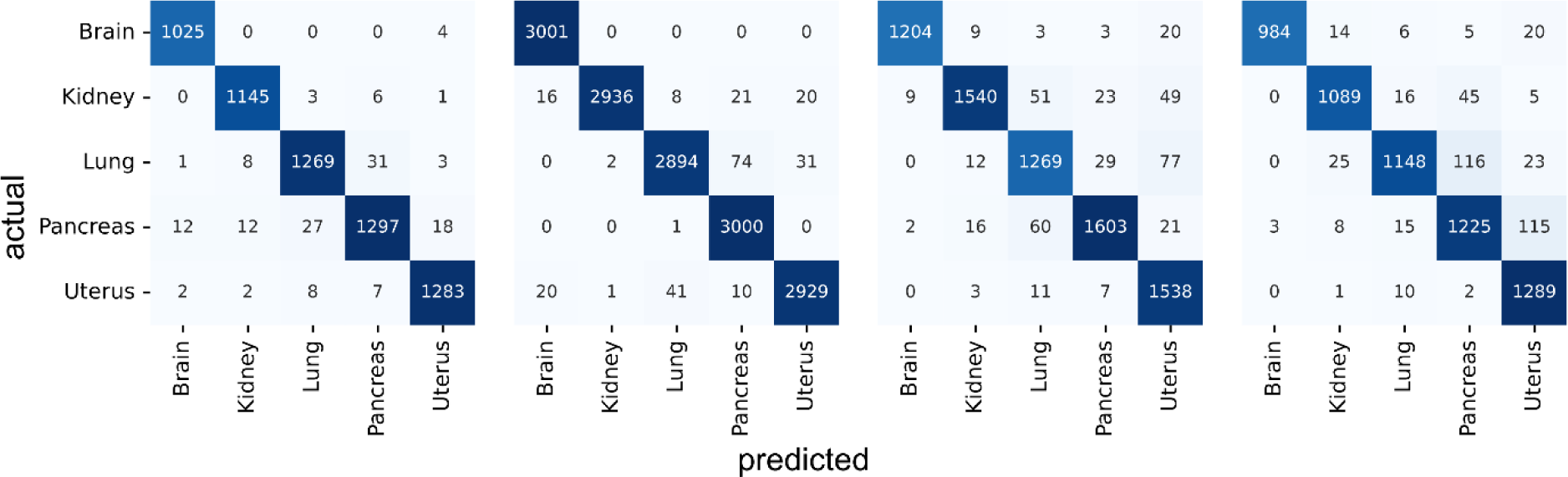
ResNet-50 confusion matrices for the tissue classification task. The confusion matrices correspond to the different scenarios presented in Table 2. From left to right, the two first panels show the test confusion matrices when the model was trained on the real data and tested on external real data and generated data, respectively. The next two panels show the confusion matrices when the model was trained on the generated data and tested on the internal and external test sets. See Materials and Methods for further information.

**Supplementary Figure 4.**
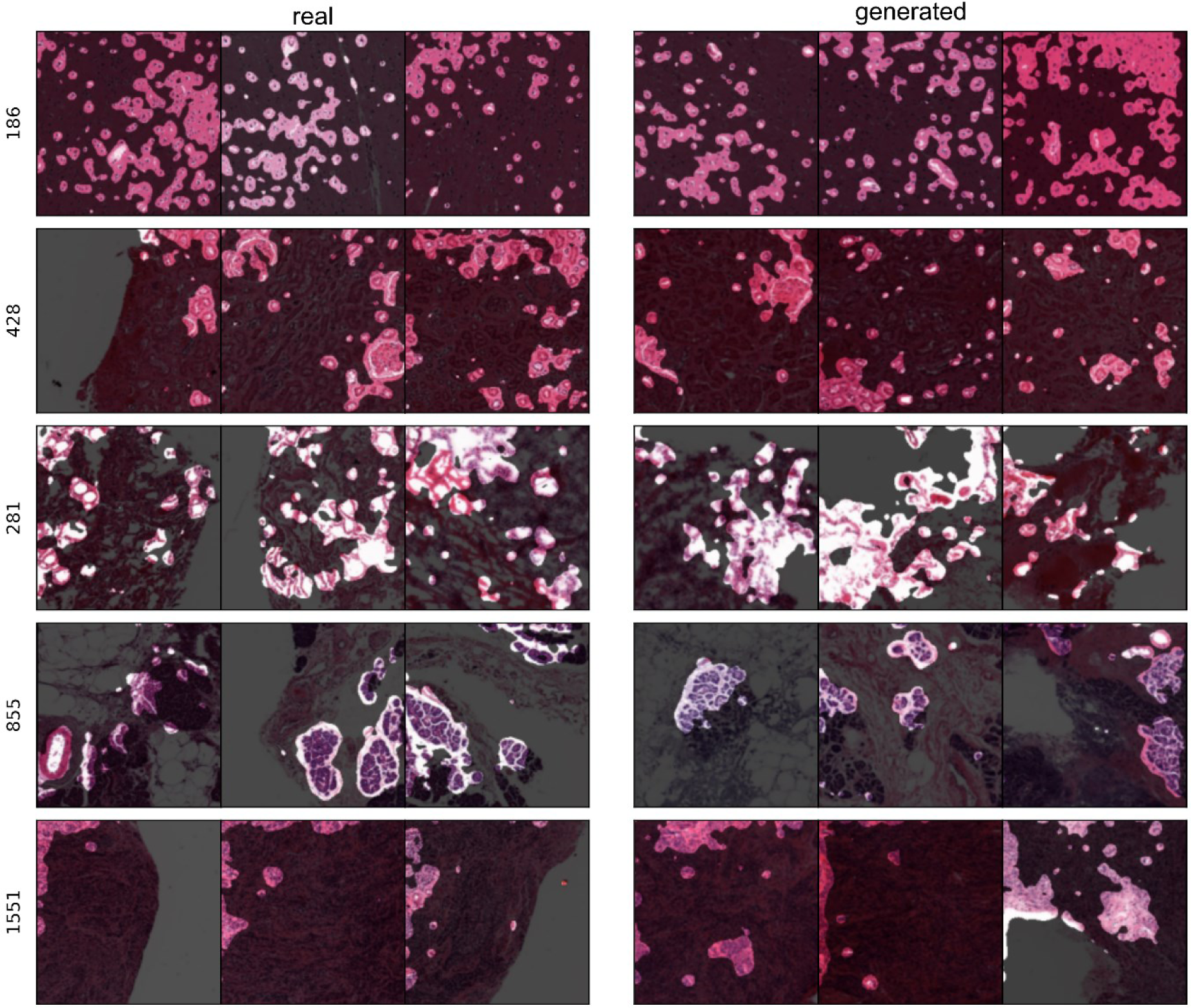
Concept relevance propagation results on the network trained with real data. The most significant concept for each tissue was chosen from the last convolutional layer of the ResNet trained on the real data, as described in the Materials and Methods section. The channel number associated with each concept is displayed on the left of each row. Three sample images from both the real data (left panel) and the generated data (right panel) were introduced into the network, and the pixels responsible for activating the concept are visually emphasized. From top to bottom, the tissues include brain, kidney, lung, pancreas, and uterus.

**Supplementary Figure 5.**
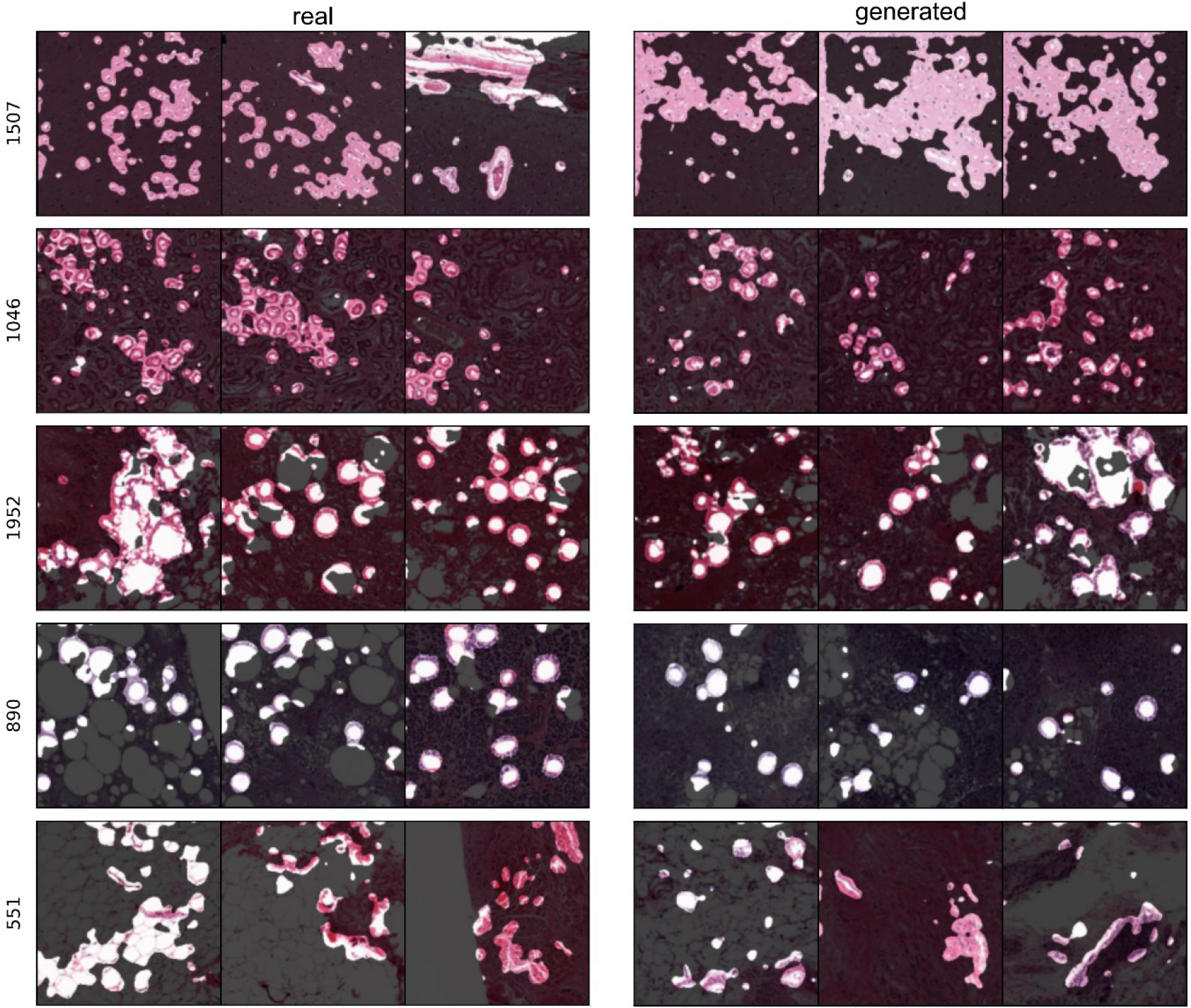
Concept relevance propagation results on the network trained with generated data. As in Supp. Figure 4, each row corresponds to the most prominent concept in the last convolutional layer of the network trained on the generated data, as indicated by the channel number on the left, specific to a particular tissue. These concepts were selected as described in the Materials and Methods section. Three sample images from the real data (left panel) and the generated data (right panel) are fed into the network, and the pixels that activate the concept are highlighted. The tissues represented from top to bottom include brain, kidney, lung, pancreas, and uterus.

**Supplementary Figure 6.**
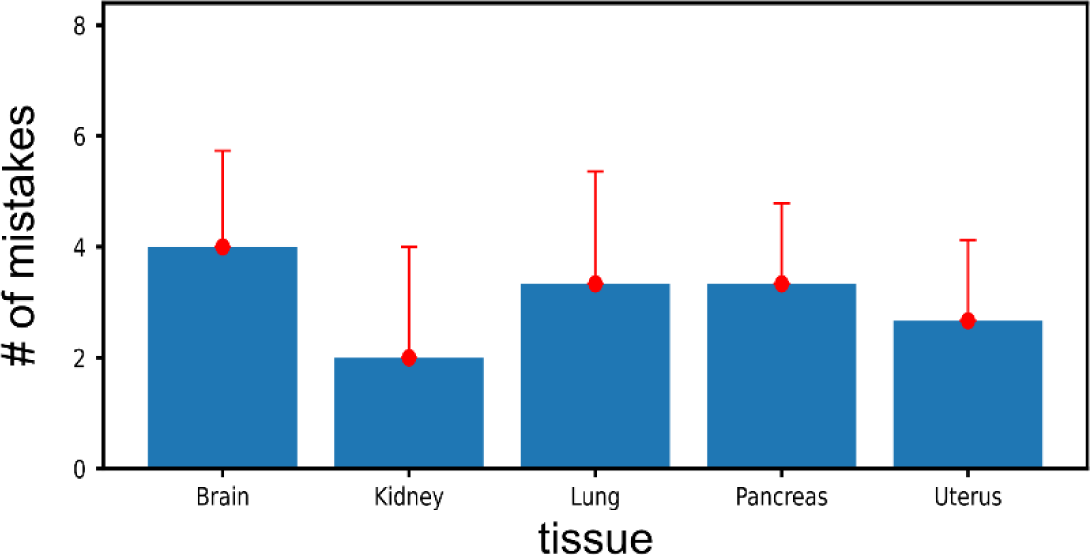
The average number of mistakes made by pathologists in discriminating generated tiles from real tiles across tissues. The mean ± standard error of the mean of the number of times that pathologists mistook a generated tile with a real tile is depicted for each tissue. The brain has the highest number of mistakes while the kidney has the lowest. The means are across pathologists.

**Supplementary Table 1.**
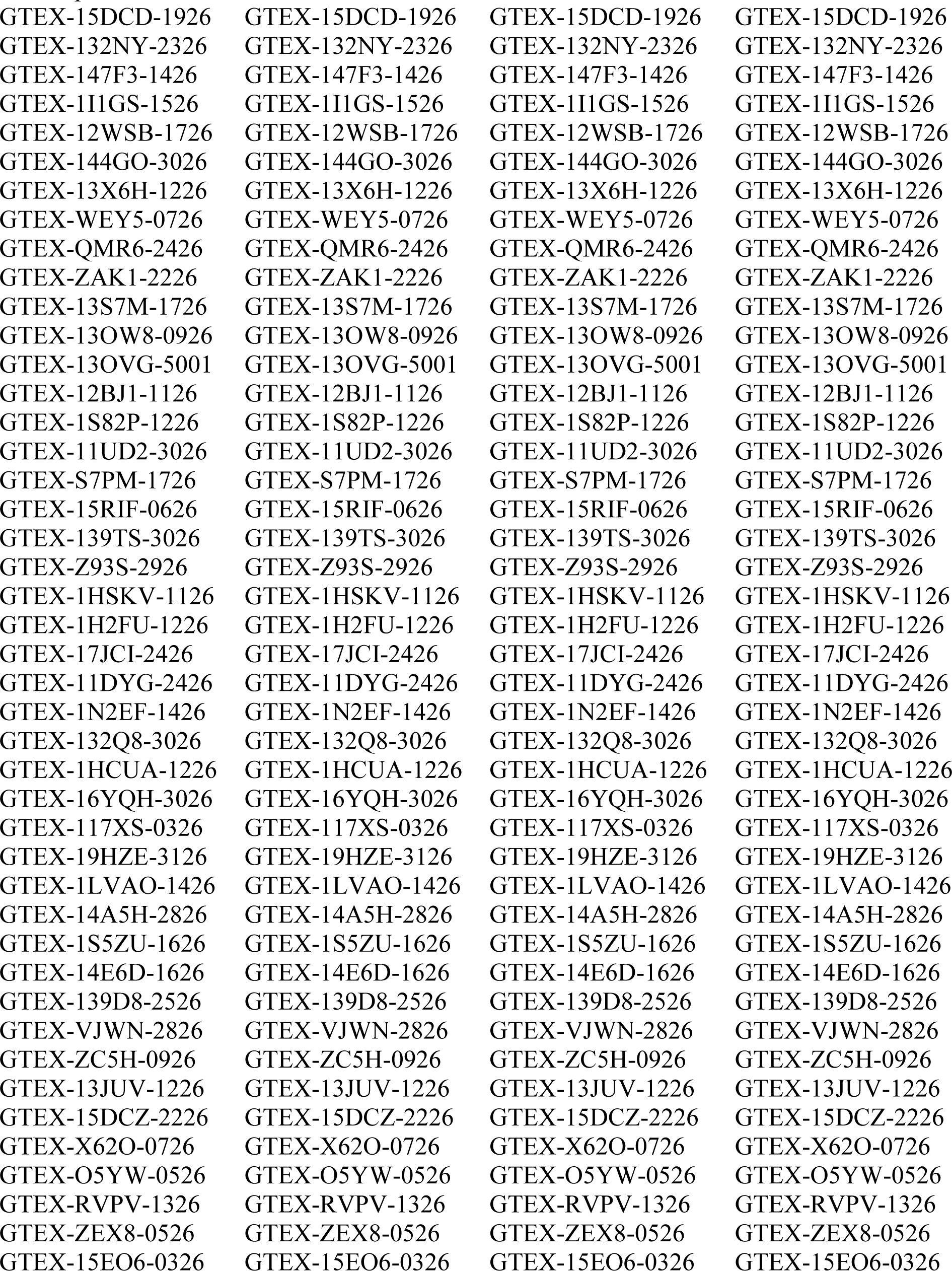
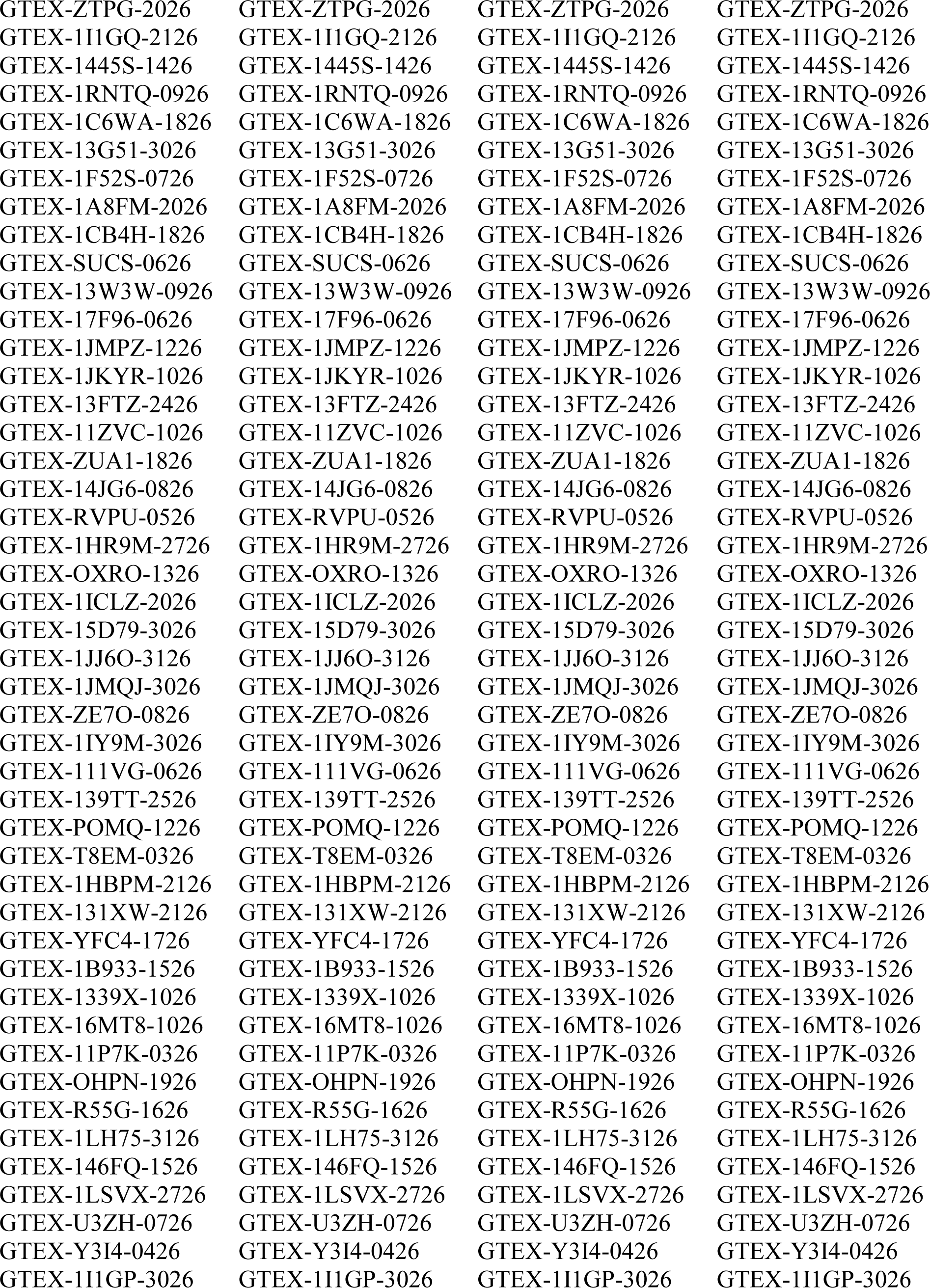
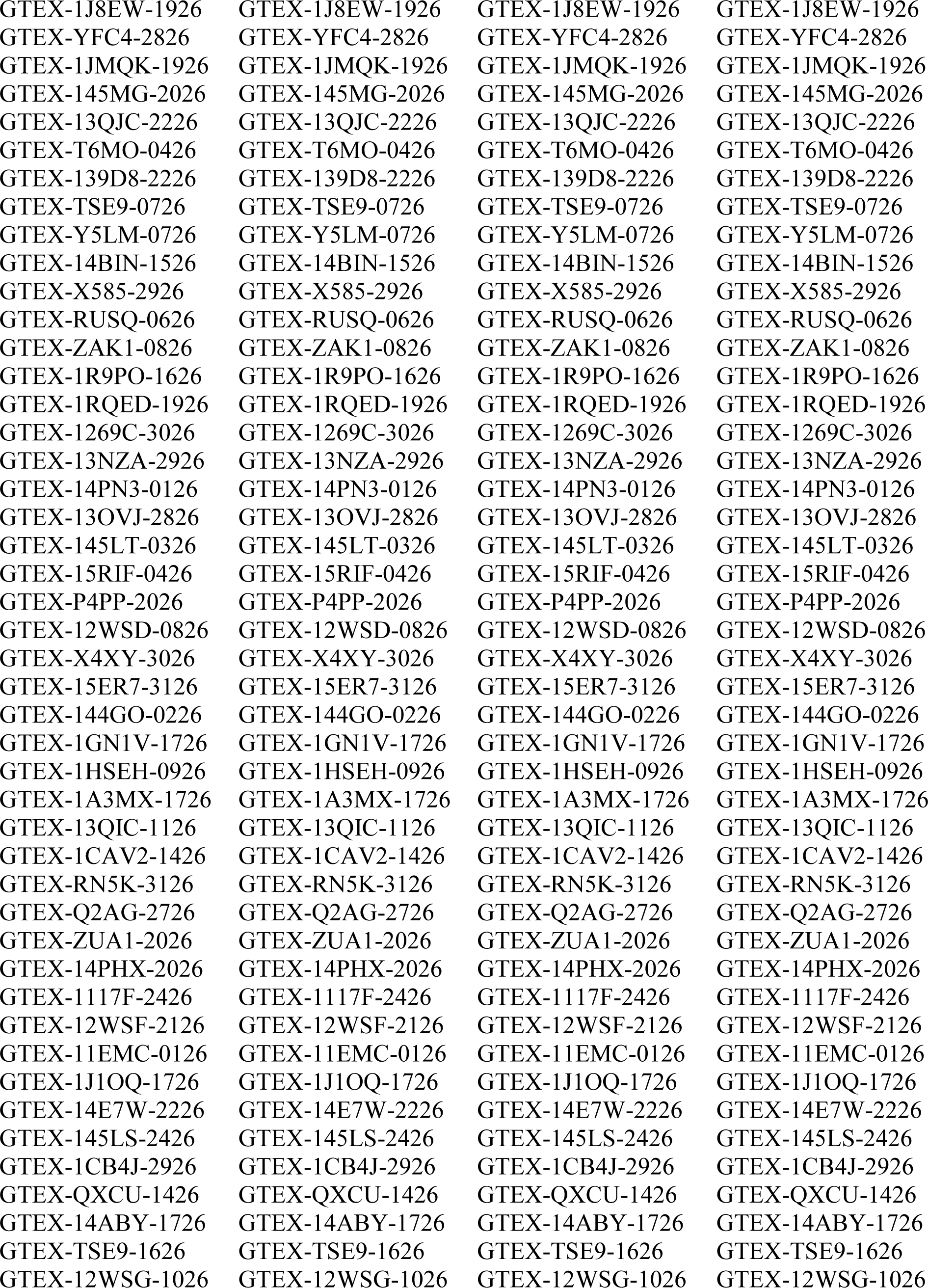
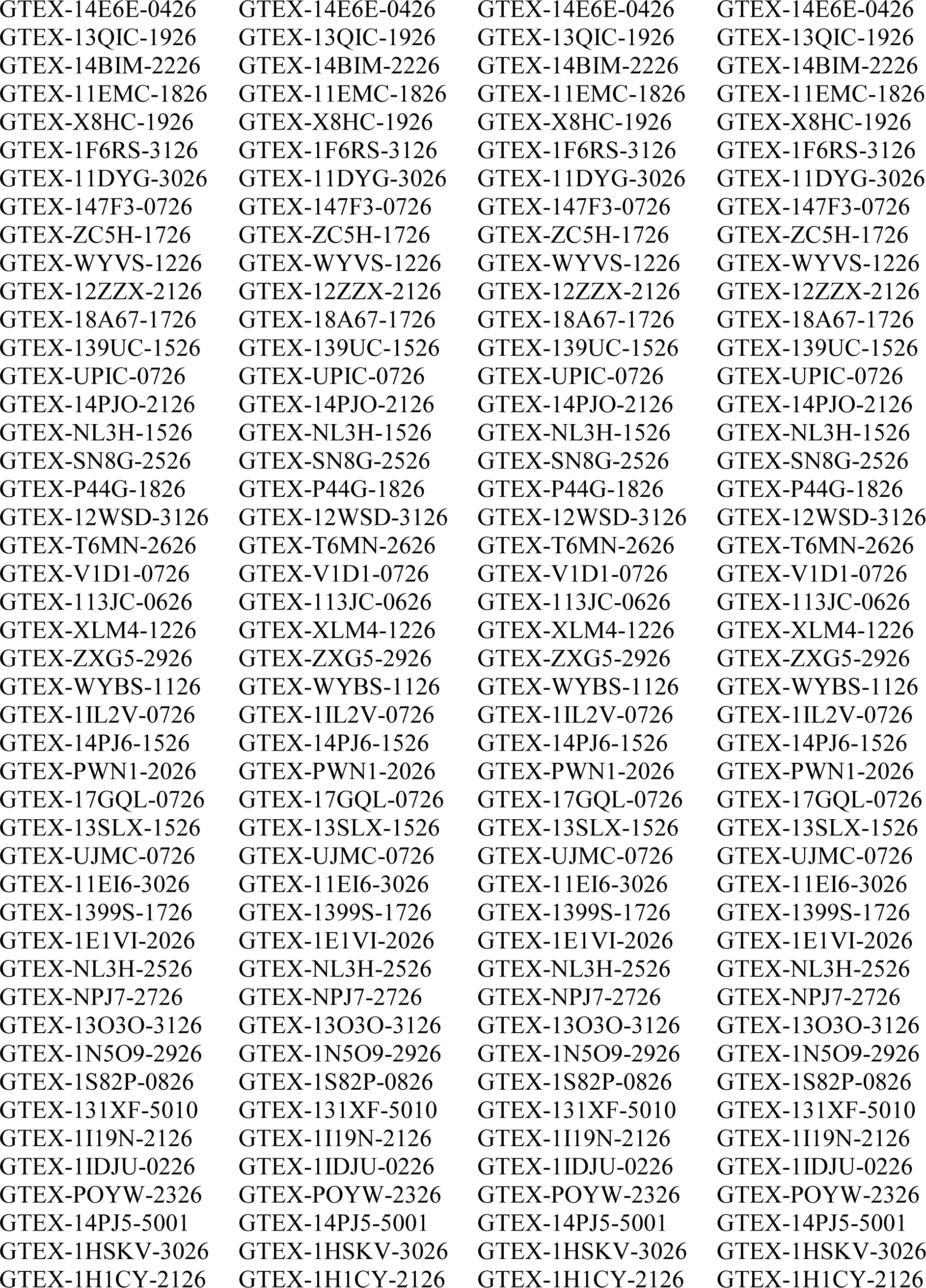
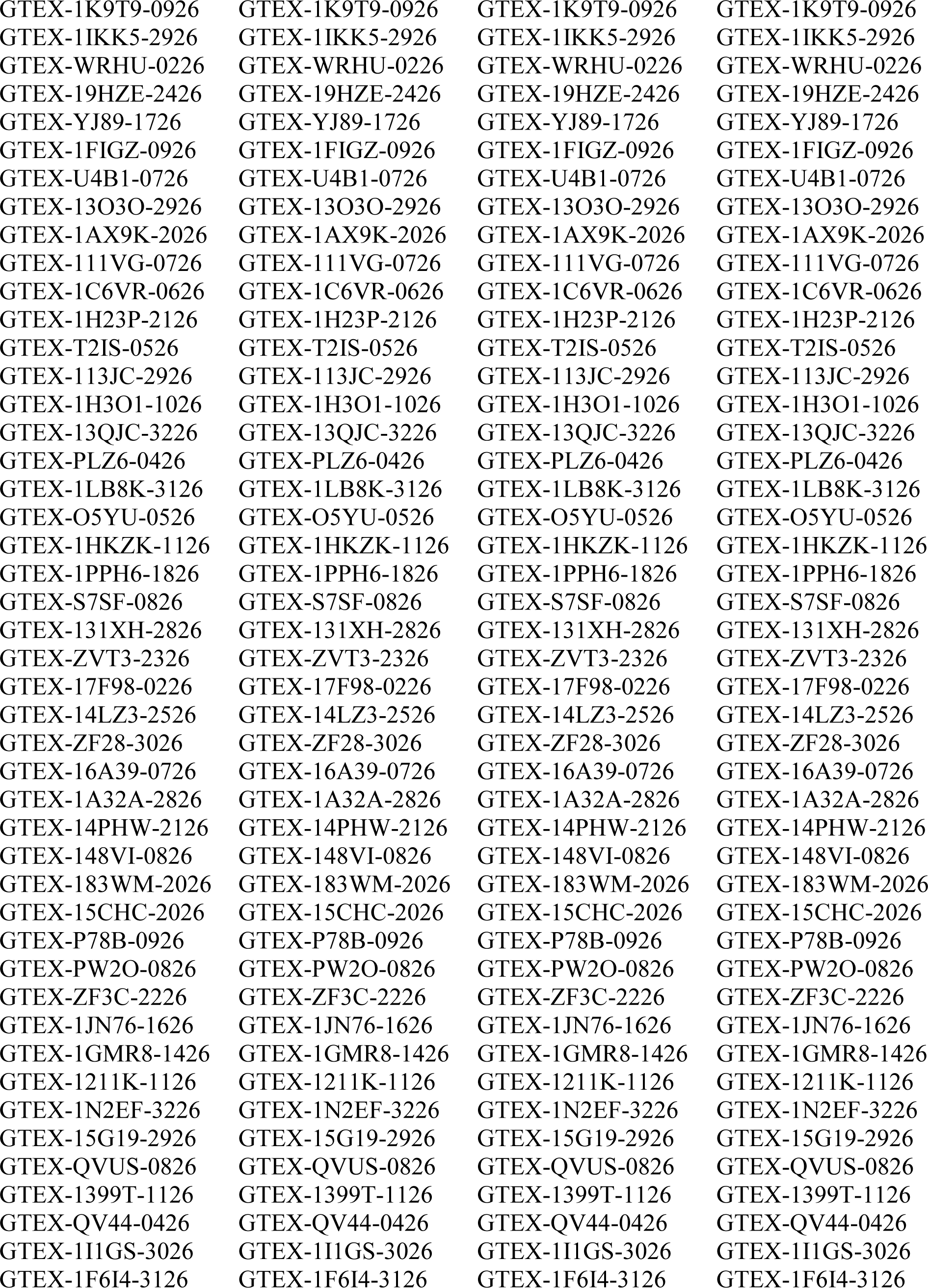
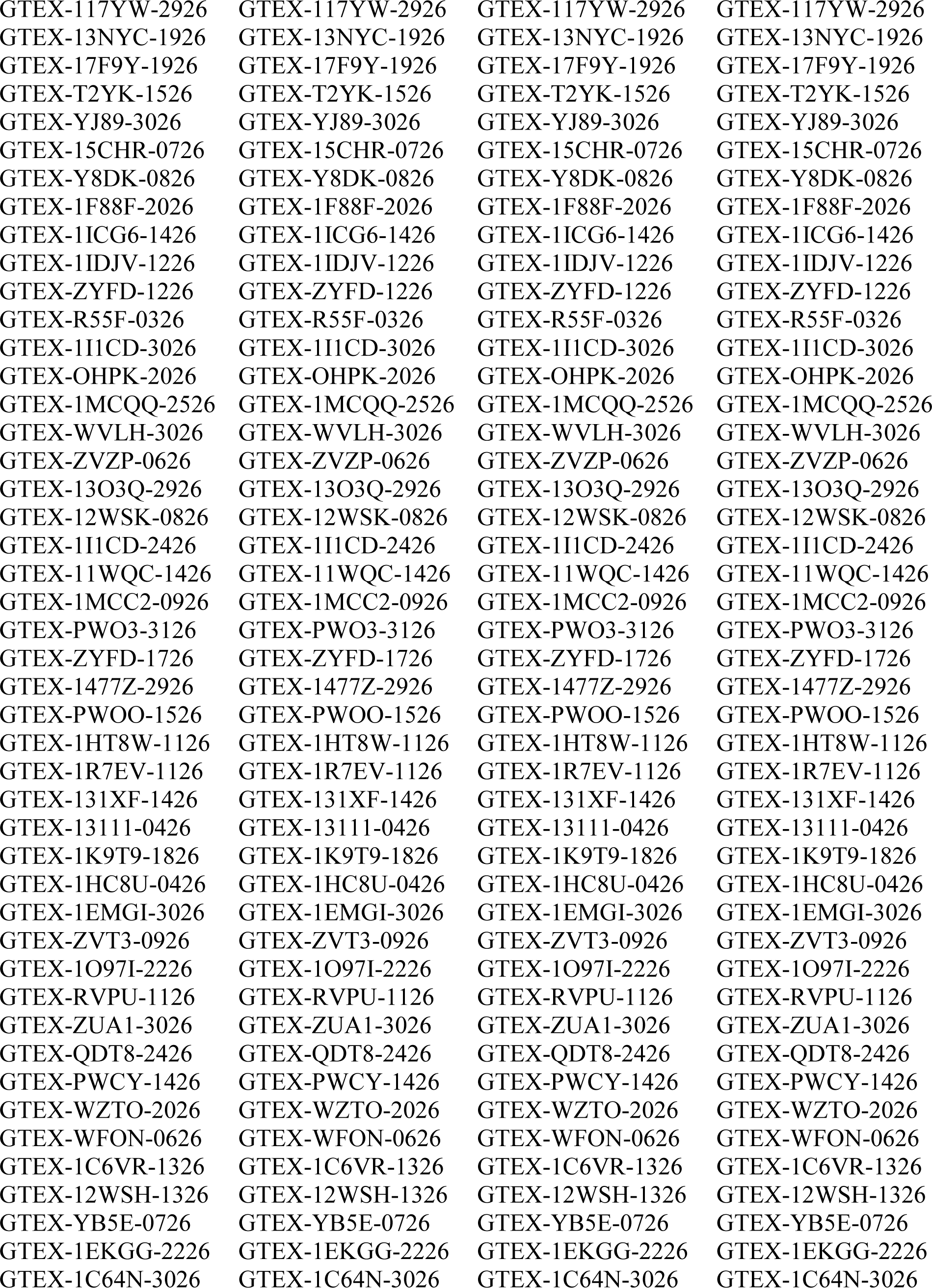
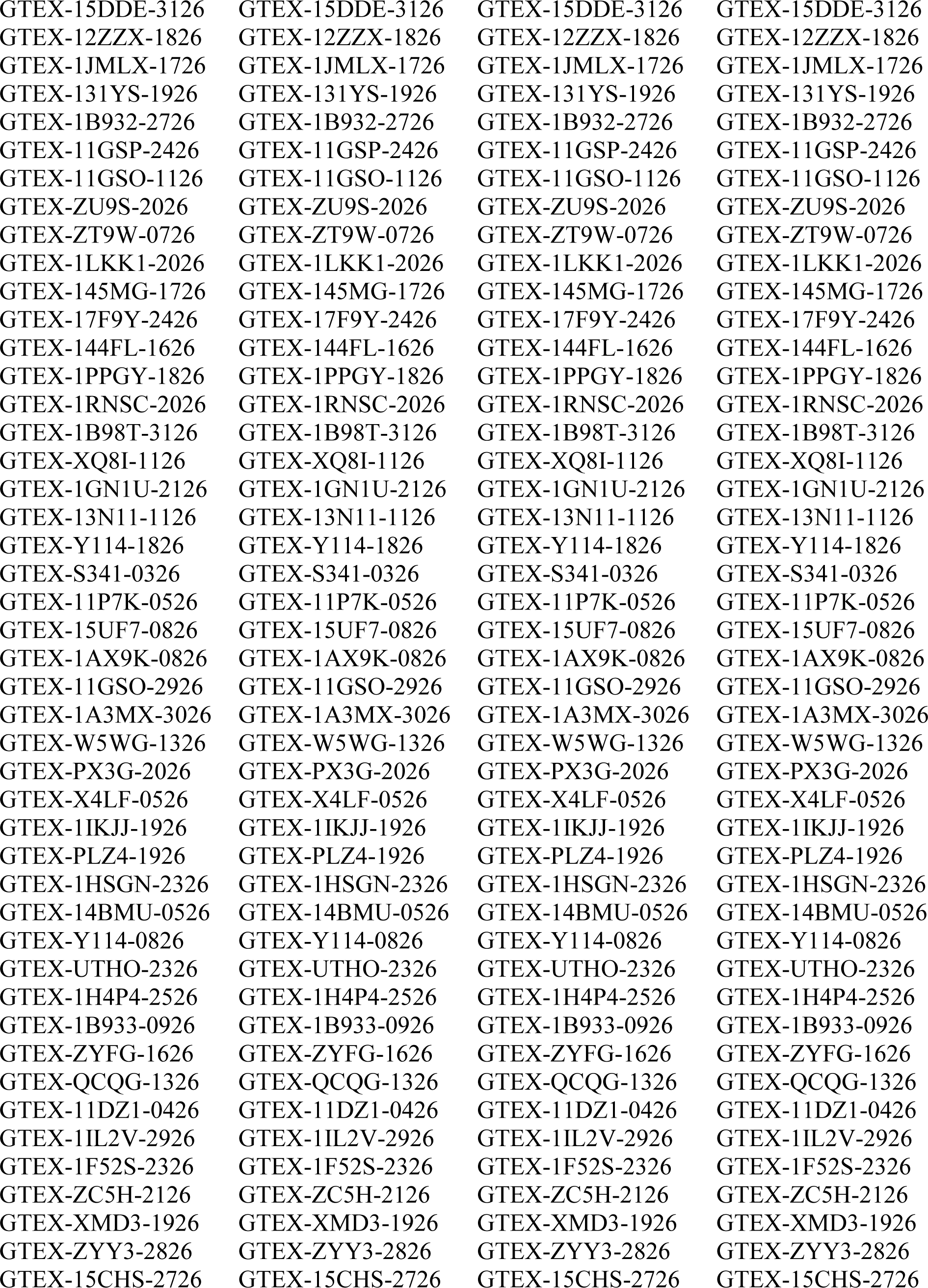
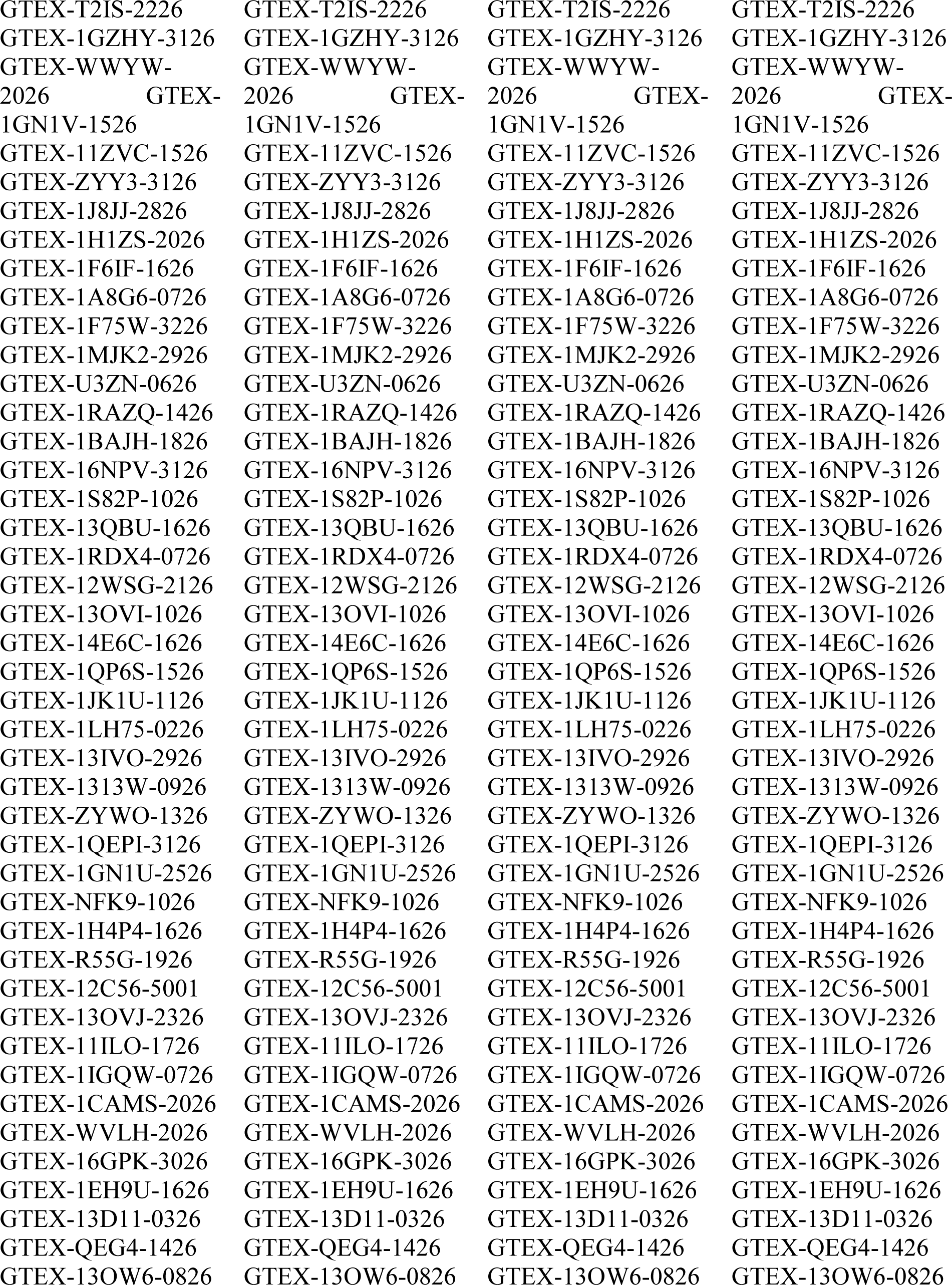
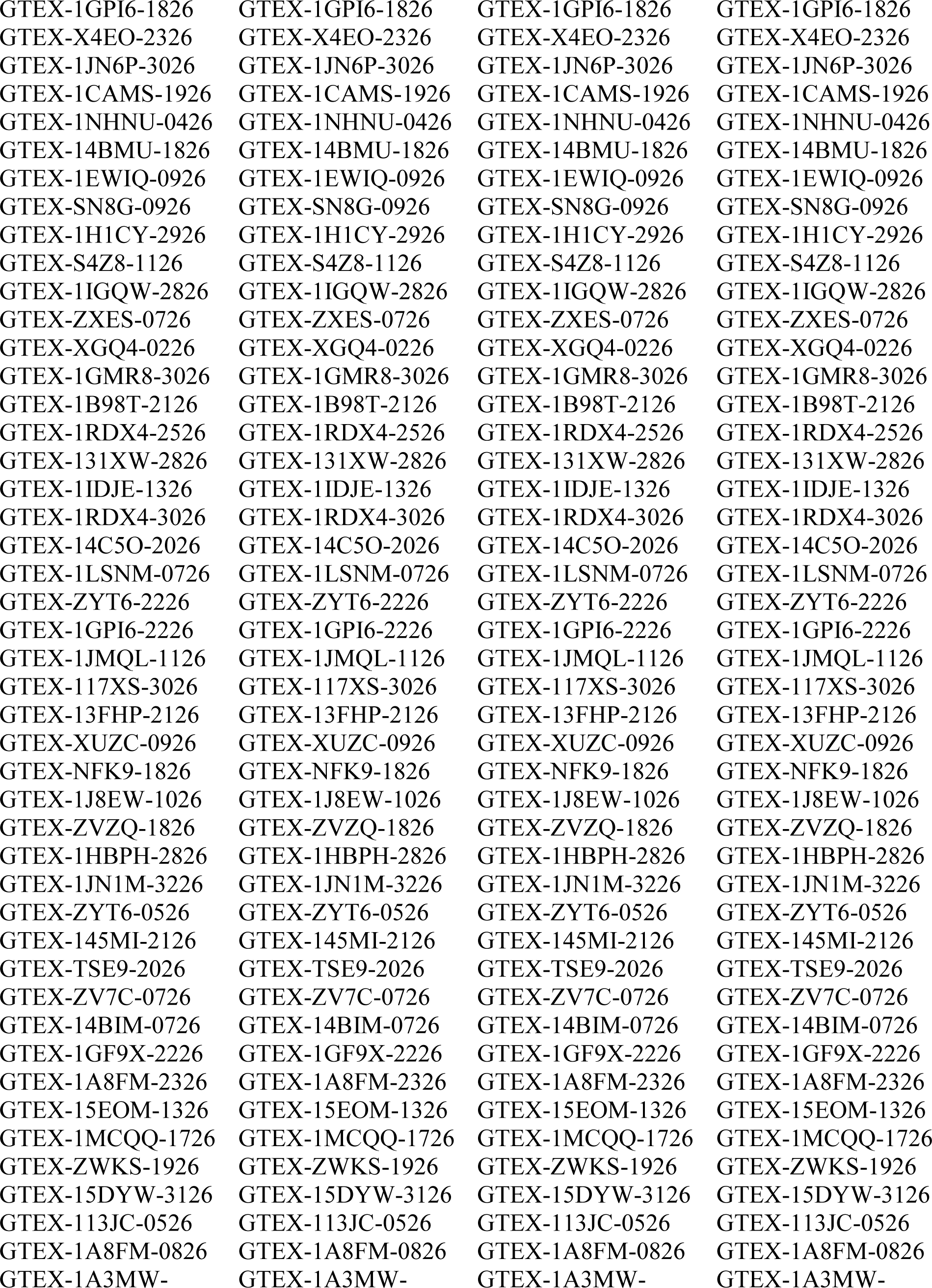
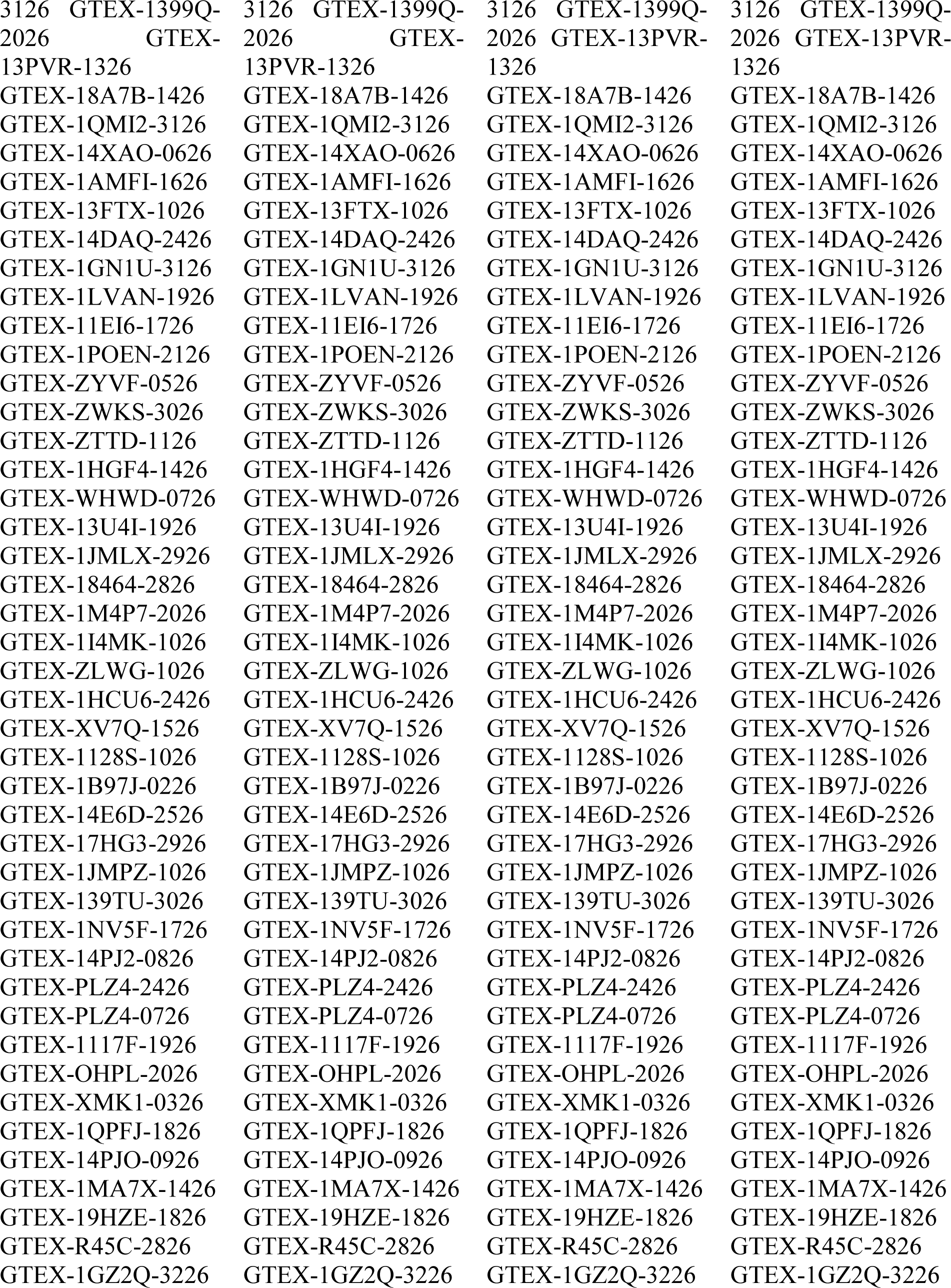
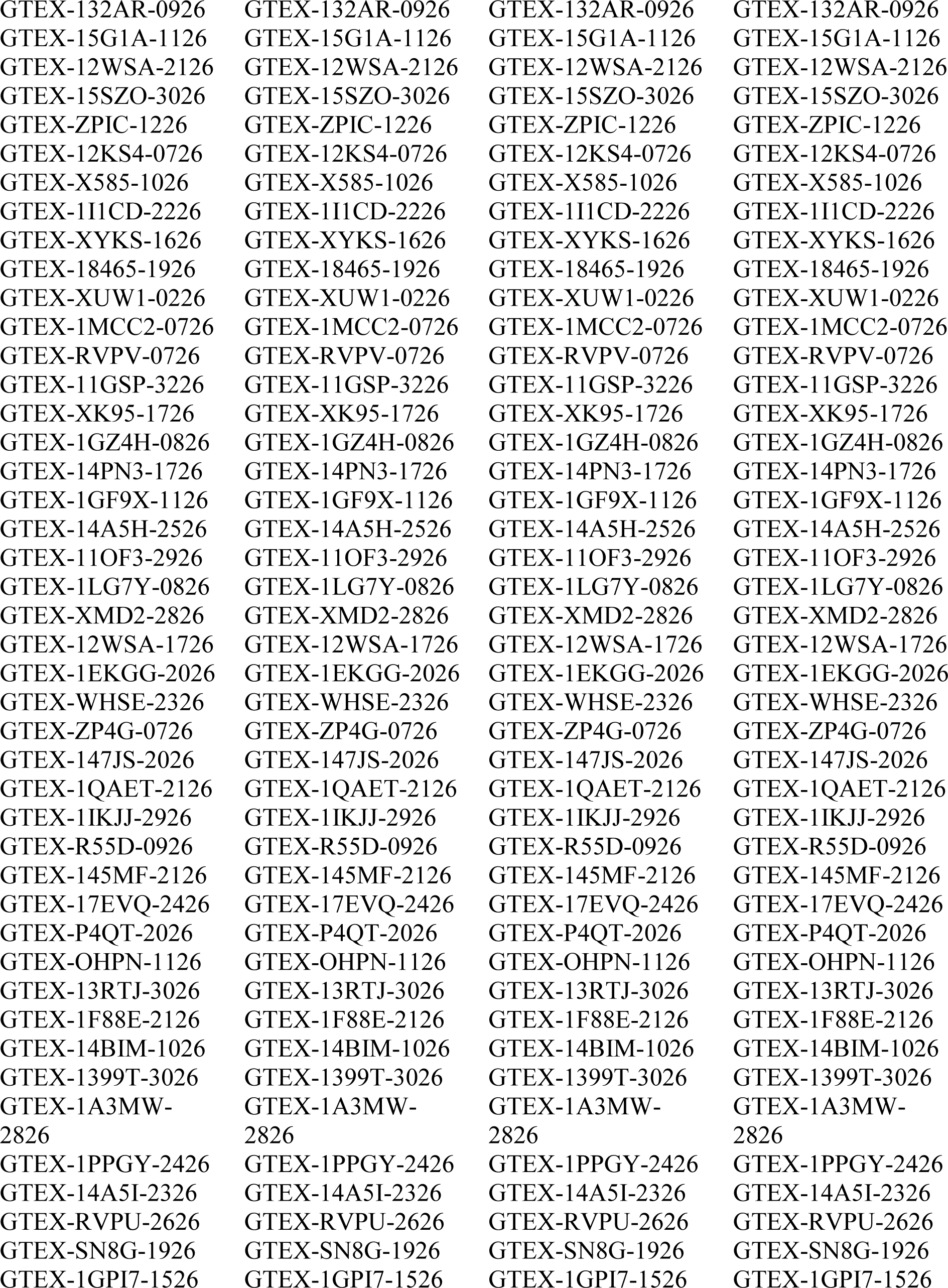
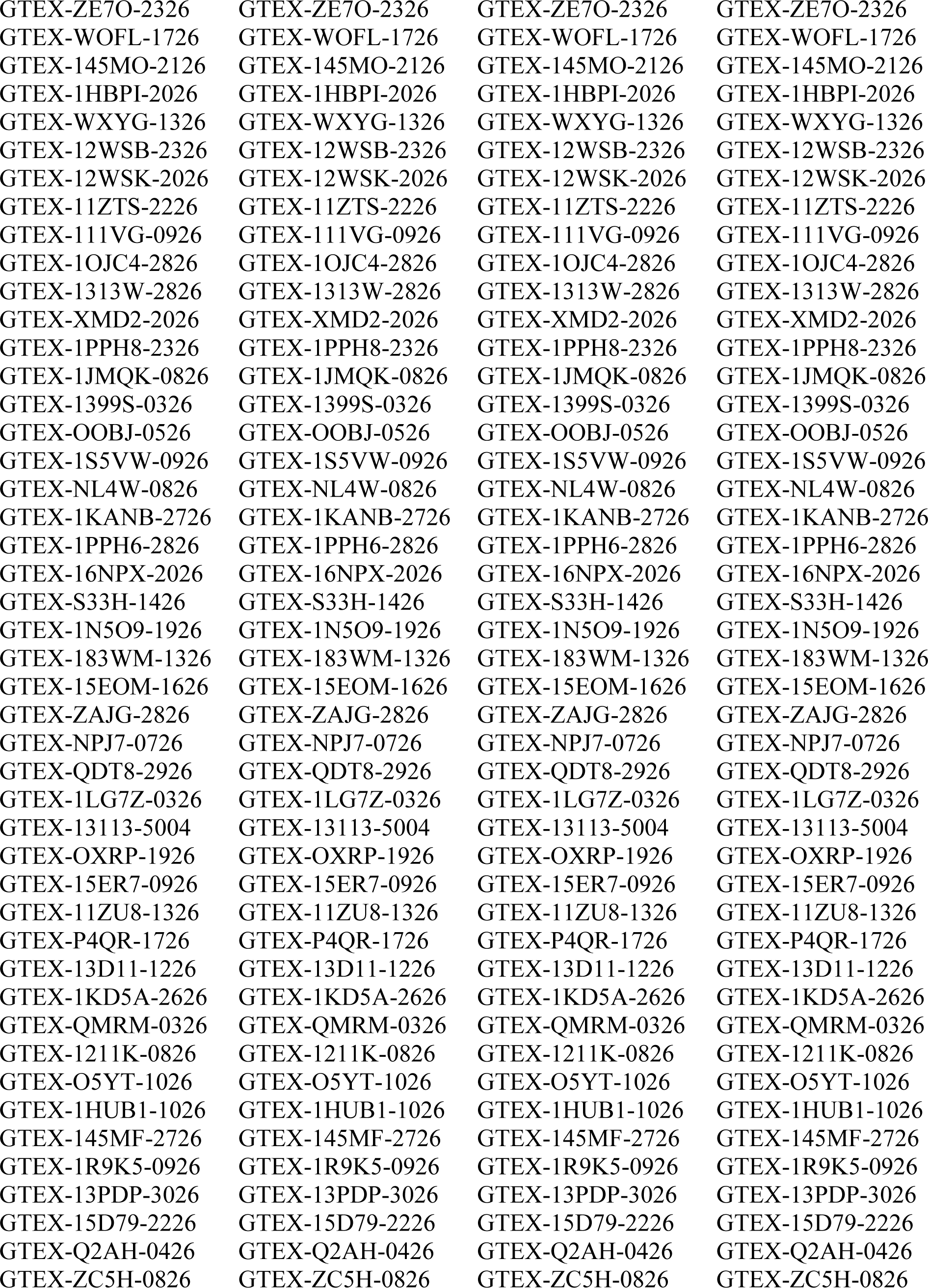
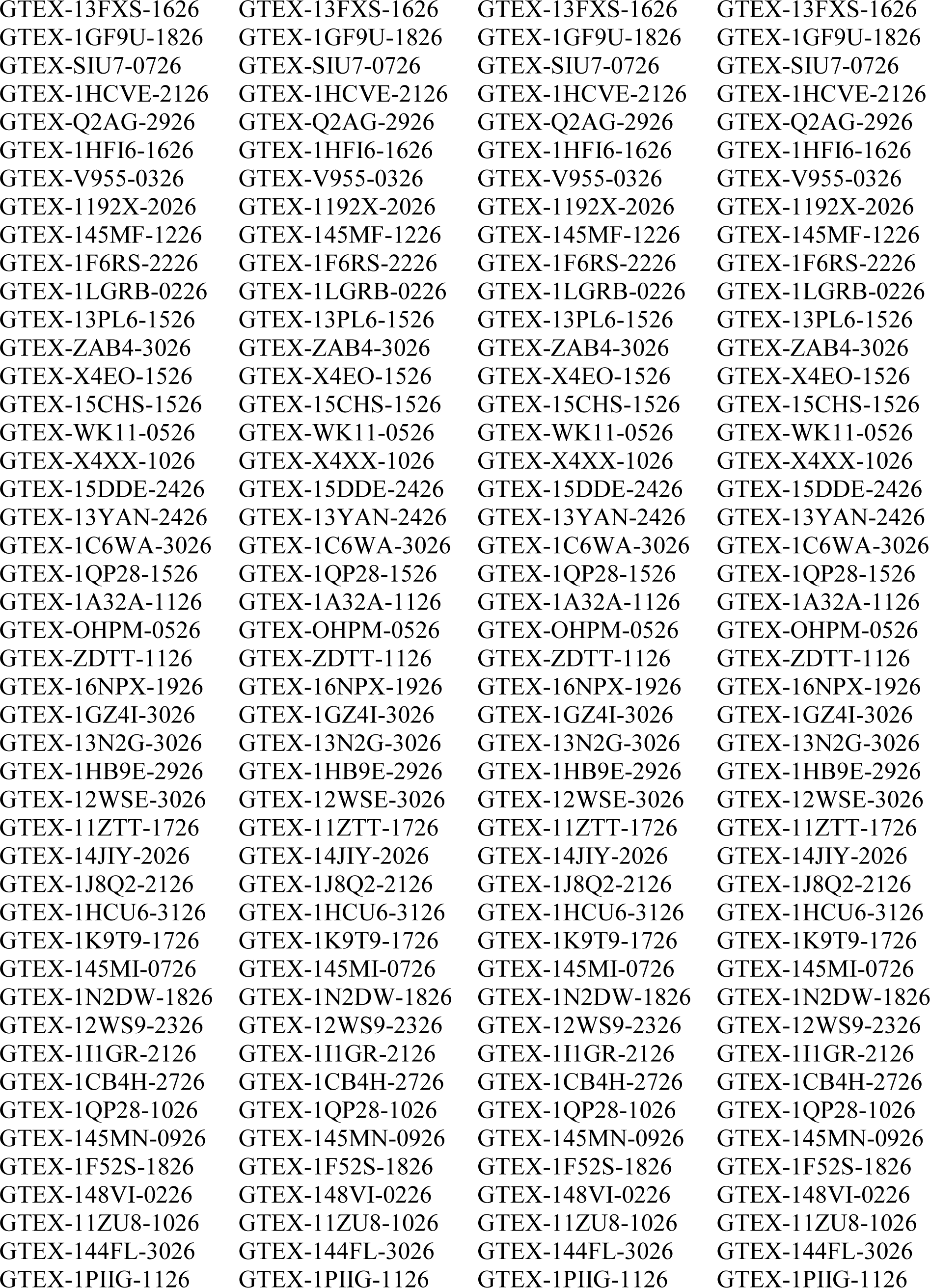
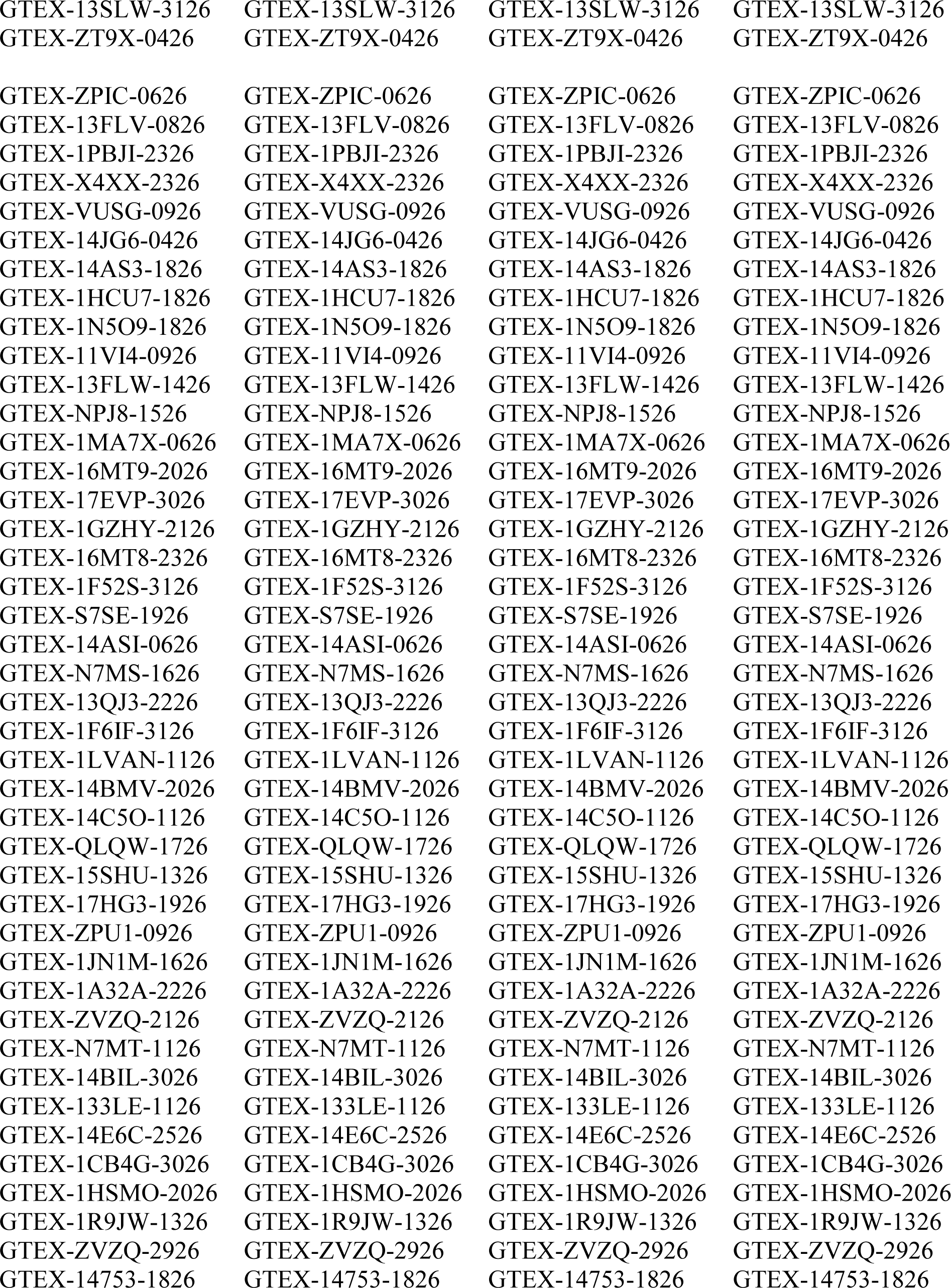
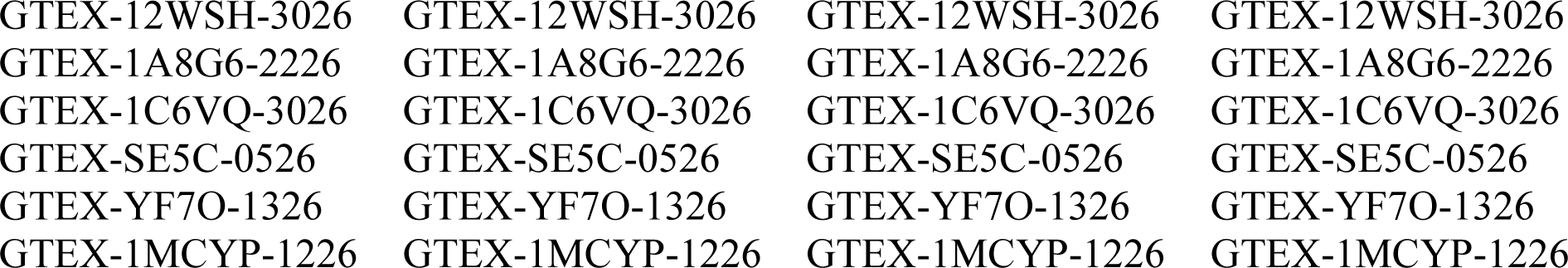
List of all the downloaded slides. The complete list of the downloaded slides is presented.

**Supplementary Table 2.** Summary of data collected. The complete list of the data used for training and testing is presented.

| **Train set** |  |  |  |  |
| --- | --- | --- | --- | --- |
| **Tissue** | **Number of tiles** | **Number of slides** | **Tiles per slide** | **Number of subjects** |
| **Brain** | 9862 | 118 | 83 | 118 |
| **Kidney** | 10848 | 116 | 93 | 116 |
| **Lung** | 10174 | 117 | 87 | 117 |
| **Pancreas** | 12807 | 119 | 107 | 119 |
| **Uterus** | 12444 | 119 | 104 | 118 |
|  | 56135 | 589 |  |  |
| **Test set** |  |  |  |  |
| **Tissue** | **Number of tiles** | **Number of slides** | **Tiles per slide** | **Number of subjects** |
| **Brain** | 1029 | 13 | 79 | 13 |
| **Kidney** | 1155 | 13 | 89 | 13 |
| **Lung** | 1312 | 13 | 101 | 13 |
| **Pancreas** | 1366 | 10 | 136 | 10 |
| **Uterus** | 1302 | 10 | 130 | 10 |
|  | 6164 | 59 |  | 59 |

**Supplementary Table 3.** Follow-up questionnaire questions. The table presents the list of questions that were asked to the pathologists in the follow-up questionnaire.

| Question |
| --- |
| To what extent did you consider color features or chromatic attributes of the images (such as intensity, value, or saliency of color) in your decisions? |
| To what extent did you consider visual quality attributes (such as blurriness, distortions, noise, etc.) in your decisions? |
| To what extent did you consider the presence of imaging artifacts or other uncommon structures in the images (such as oversaturation of color, dark spots, edges, white spaces, etc.) in your decisions? |
| To what extent did you consider the presence of unrealistic tissue morphologies in the images (e.g., mixed brain/kidney morphologies in one image) in your decisions? |
| To what extent did you consider unrealistic biological features of the images (such as non-biological cell shapes or unrealistic cell concentration) in your decisions? |
| To what extent did you consider cell nuclei shape, abundance, or proportion? |
| To what extent did you evaluate the magnification level of the tiles? |

**Supplementary Table 4.** Comparison of quantitative evaluation metrics with other studies. This table compares the quantitative metrics obtained in our study with those reported in other studies utilizing diffusion models for generating digital pathology images. To ensure a fair comparison, we recalculated the precision and recall using features extracted from an ImageNet pretrained ResNet50 network. Additionally, we provide information on the number of tissues (\# of tissues) in each respective study to facilitate a comprehensive comparison of the results. IS represents Inception Score, while FID stands for Fréchet Inception Distance. The direction of the arrow in parentheses indicates whether higher or lower values are preferred. Bolded values show the best obtained results.

| Study | Number of Tissues | IS (↑) | FID (↓) | Precision (↑) | Recall (↑) |
| --- | --- | --- | --- | --- | --- |
| Current | 5 | **2.91** | 35.11 | **0.82** | 0.40 |
| Moghadam et al. | 1 | 2.08 | 20.11 | 0.25 | **0.85** |
| Shrivastava et al. x20 | 1 | 2.7 | 15.7 | - | - |
| Shrivastava et al. x10 | 1 | 2.3 | 38.1 | - | - |
| Xu et al. dataset 1 | 1 | - | 12.4 | 0.6 | 0.4 |
| Xu et al. dataset 2 | 1 | - | 13.39 | 0.6 | 0.44 |
| Xu et al. dataset 3 | 1 | - | 36.18 | 0.51 | 0.5 |

**Supplementary Table 5.** Global explainability analyses. The first column contains a channel (concept) related to the identification of the class listed in the second column. Columns 3 and 4 display the class ID and its corresponding normalized relevance obtained from both the entire synthetic and real datasets. This information is provided for both the model trained on real data and the model trained on generated data.

| **Model trained on real data** |  |  |  |
| --- | --- | --- | --- |
| **Channel** | **Tissue (id)** | **Classes (Norm Rel)**<br>**generated data** | **Classes (Norm Rel)**<br>**real data** |
| 186 | Brain (0) | **0** (1), 4 (0.14), 3 (0.08), 2 (0.073), 1 (0.036) | **0** (1), 2 (0.23), 3 (0.17), 4 (0.13), 1 (0.058) |
| 428 | Kidney (1) | **1** (1), 0 (0.176), 2 (0.13), 3 (0.02), 4 (0.02) | **1** (1), 2 (0.22), 0 (0.21), 4 (0.037), 3 (0.026) |
| 281 | Lung (2) | **2** (1), 4 (0.3), 3 (0.085), 1 (0.048), 0 (-0.0021) | **2** (1), 4 (0.37), 0 (0.15), 3 (0.13), 1 (0.09) |
| 327 | Pancreas (3) | **3** (1), 0 (0.58), 1 (0.33), 2 (0.19), 4 (0.008) | **3** (1), 0 (0.35), 1 (0.29), 2 (0.20), 4 (0.16) |
| 1551 | Uterus (4) | **4** (1), 1 (0.083), 2 (0.079), 0 (0.016), 3 (0.004) | **4** (1), 0 (0.147), 3 (0.090), 2 (0.083), 1 (0.068) |
| **Model trained on generated data** |  |  |  |
| **Channel** | **Tissue (id)** | **Classes (Norm Rel)**<br>**generated data** | **Classes (Norm Rel)**<br>**real data** |
| 1507 | Brain (0) | **0** (1.00), 1 (0.10), 2 (0.02), 3 (0.00), 4 (0.00) | **0** (1.00), 3 (0.46), 1 (0.10), 4 (0.03), 2 (0.01) |
| 1046 | Kidney (1) | **1** (1.00), 3 (0.25), 4 (0.15), 0 (0.11), 2 (0.06) | **1** (1.00), 4 (0.49), 3 (0.25), 0 (0.11), 2 (0.06) |
| 1952 | Lung (2) | **2** (1.00), 3 (0.48), 1 (0.03), 0 (0.00), 4 (0.00) | **2** (1.00), 3 (0.49), 4 (0.20), 1 (0.03), 0 (0.00) |
| 890 | Pancreas (3) | **3** (1.00), 2 (0.95), 4 (0.00), 1 (0.00), 0 (0.00) | **3** (1.00), 2 (0.94), 4 (0.14), 0 (0.00), 1 (0.00) |
| 551 | Uterus (4) | **4** (1.00), 2 (0.00), 1 (0.00), 3 (0.00), 0 (0.00) | **4** (1.00), 2 (0.05), 0 (0.03), 1 (0.02), 3 (0.02) |

## Footnotes

1 https://github.com/histolab/histolab

2 https://github.com/lucidrains/denoising-diffusion-pytorch/tree/main

3 https://torchmetrics.readthedocs.io/en/stable/

4 https://github.com/clovaai/generative-evaluation-prdc

5 https://pytorch.org/

6 https://www.pytorchlightning.ai/index.html

7 https://github.com/rachtibat/zennit-crp

8 https://chat.openai.com/

## Notes

### Competing Interest Statement

The authors have declared no competing interest.

### Author Declarations

The study used ONLY openly available human data that were originally located at: https://gtexportal.org/home/

